# Unanticipated Global Emergence of Multiple *Pneumocystis jirovecii* Mutants Selected by Mycophenolic Acid Driving Increasing Outbreaks in Solid Organ Transplant Recipients

**DOI:** 10.1101/2025.06.17.25328583

**Authors:** Liang Ma, Ming Hao, Weizhong Chang, Xilong Deng, Junfeng Sun, Marwan M. Azar, Alexia Cusini, Thomas Fehr, Sara Gianella, Norihiko Goto, Jannik Helweg-Larsen, Grace Handley, Cedric Hirzel, Laurence Huang, Regina Konrad, Nicolas J. Mueller, Shinichi Oka, Lingai Pan, Li Peng, Andreas A. Rostved, Monica Sassi, Andreas Sing, Benjamin Spielman, Laura F. Walsh, Yubao Wang, Hirohisa Yazaki, Lizbeth Hedstrom, Tomozumi Imamichi, Joseph A. Kovacs

**Author notes:** Correspondence to: Dr. Liang Ma and Dr. Joseph A. Kovacs, Critical Care Medicine Department, National Institutes of Health Clinical Center, Bethesda, MD 20892, USA.

## Abstract

**Background:** Classified by the WHO as one of the 19 most dangerous fungal pathogens, *Pneumocystis jirovecii* has been associated with increasing outbreaks of *Pneumocystis* pneumonia (PCP) among solid organ transplant (SOT) recipients worldwide. Mycophenolic acid (MPA), an inosine monophosphate dehydrogenase (IMPDH) inhibitor commonly used as an immunosuppressant to prevent organ rejection, is a risk factor for PCP. However, MPA also displays antifungal activity, potentially protecting against PCP, despite not being used to treat it. Therefore the underlying factors driving these outbreaks remain undefined.

**Methods:** In this international multicenter retrospective observational study, *P. jirovecii* samples were collected from 96 SOT patients (including 94 from nine separate outbreaks and 84 on MPA therapy) and 67 non-transplant controls (none on MPA), between 1986 and 2020 across six countries in Europe, North America and Asia. All samples underwent extensive targeted sequencing of the *P. jirovecii* inosine monophosphate dehydrogenase (*impdh*) gene and multiple genetic markers, with selected samples further analyzed for complete mitogenome and restriction fragment length polymorphisms. Computational modeling was employed to predict the effects of IMPDH mutations on protein structure and MPA binding.

**Results:** Six *impdh* mutations (including one previously reported) were identified, with frequencies of 4-21% each in SOT patients and 0-1% in controls. These mutations were strongly associated with prior MPA exposure and showed marked geographic segregation and temporal shifts. Four mutations were each linked to multiple distinct genotype profiles, representing separate *P. jirovecii* strains. Structure modeling predicted that these four mutations reduced protein stability and binding affinity to MPA.

**Conclusions:** This study suggests that the widespread use of MPA in SOT recipients has unexpectedly driven the emergence of multiple *impdh* mutations in *P. jirovecii*, each presumably arising independently in multiple strains worldwide. These mutations likely confer drug resistance and provide a selective survival advantage to *P. jirovecii* in SOT recipients exposed to MPA, thereby facilitating transmission and outbreaks. These findings have significant implications for the prevention and clinical management of PCP in SOT recipients, highlighting a rare example of how antimicrobial resistance can emerge through unexpected pathways, transcending conventional antimicrobial use and emphasizing the need for increased vigilance and strategic adaptation in clinical practice.

## Introduction

Once as a hallmark of HIV/AIDS, the incidence of *Pneumocystis* pneumonia (PCP) has declined significantly in this patient population, primarily due to the widespread implementation of combination antiretroviral therapy (cART) as well as the broad utilization of anti-*Pneumocystis* prophylaxis. However, over the past two decades, the incidence of PCP has been steadily increasing in non-HIV immunocompromised populations.^1,2^ The growing public health importance of *Pneumocystis* has led the World Health Organization (WHO) to include it among the 19 most dangerous fungal pathogens.^3^ A recent reassessment of this list has recommended elevating *Pneumocystis* to the Critical Priority Group, alongside *Aspergillus fumigatus*, *Candida* spp., and *Cryptococcus neoformans*,^4^ due to its substantial disease burden, with an estimated 505,000 cases and 214,000 deaths annually, as well as its evolving epidemiology and treatment challenges.^4,5^

Solid organ transplant (SOT) recipients represent one of the most prominent non-HIV populations at risk for PCP, with frequent outbreaks reported globally, particularly among renal transplant recipients. The occurrence of these outbreaks has spurred global efforts to investigate their underlying causes and potential intervention strategies.^6–12^ Numerous molecular typing studies have characterized *P. jirovecii* strains involved in the outbreaks, demonstrating either a single or a limited number of strains in each outbreak but without defining the underlying factors driving the outbreaks.

A recent study of PCP in French SOT patients has linked a nonsynonymous mutation (A261T) in the *P. jirovecii impdh* gene to prior therapy with mycophenolic acid (MPA),^13^ a commonly prescribed immunosuppressant to prevent organ rejection. Further genotyping studies using Sanger sequencing of a limited number of genetic loci showed that the A261T mutation was associated with a single *P. jirovecii* strain.^14^ These observations led to the hypothesis that the prophylactic use of MPA to prevent organ rejection may have led to the selection of a specific *P. jirovecii* strain, which preferentially circulates among SOT recipients, thus contributing to transmission and outbreaks. We were intrigued by this hypothesis and leveraged a large collection of samples from prior collaborative studies of PCP outbreaks following SOT across the USA, Europe and Asia,^6–11^ utilizing comprehensive genetic and structural modeling analyses, to evaluate whether the hypothesis was supported by data from a more geographically and temporally diverse sampling.

## Methods

### Patients and *P. jirovecii* samples

Residual DNA extracts from previous studies were utilized to analyze 155 *P. jirovecii* isolates (Table 1);^6–11^ whole genome next-generation sequencing (NGS) data were available from public database for eight isolates (Supplementary Table S3). Patients originated from ten cities across six countries, including the USA, Denmark, Germany, Switzerland, Japan, and China. All patients were confirmed to be infected with *P. jirovecii* by microscopical detection of stained respiratory samples and/or PCR or NGS detection of *P. jirovecii* DNA. The patients were divided into two groups: the solid organ transplant (SOT) recipient group and the control group (Table 1).

**Table 1:**
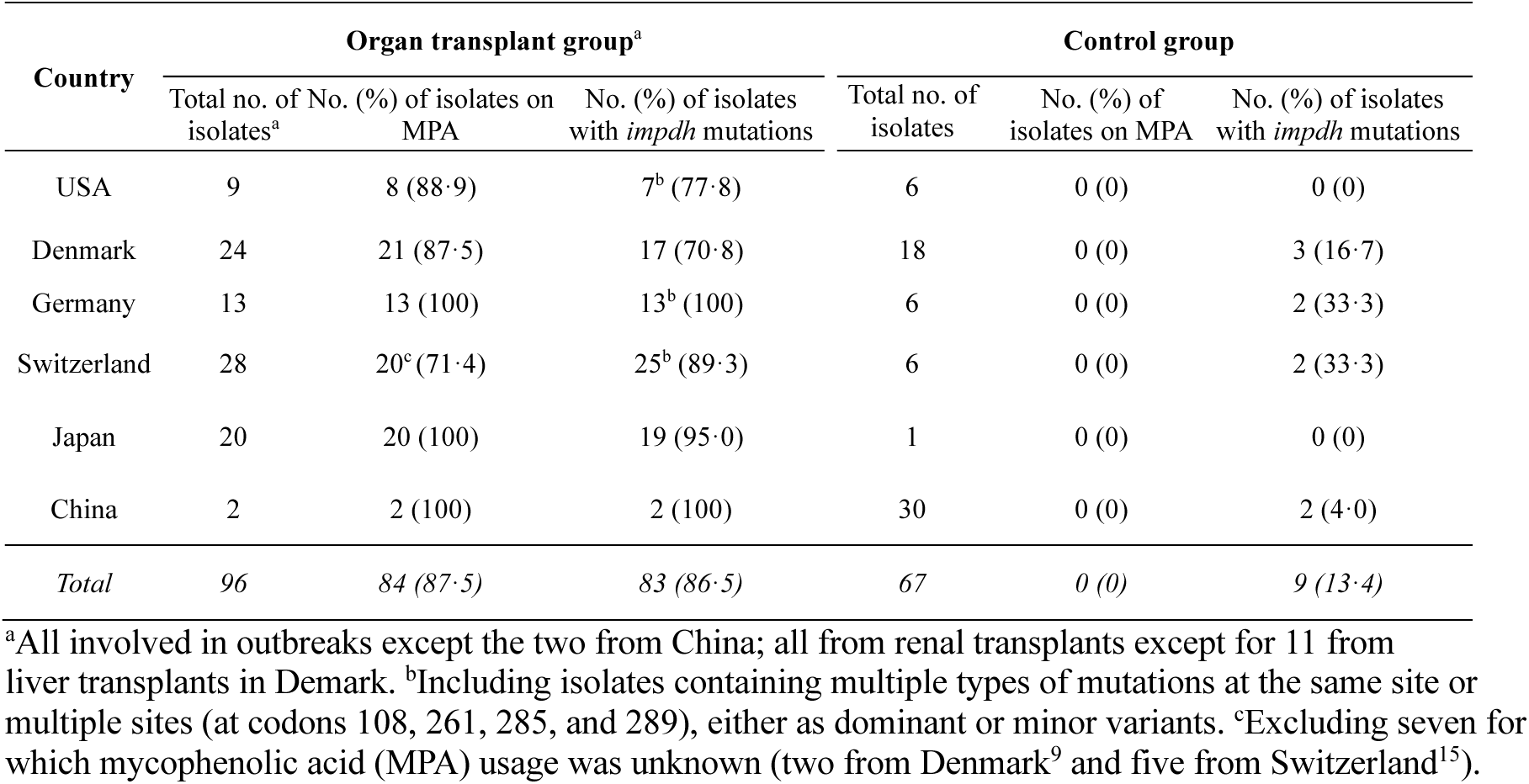
*P. jirovecii* isolates and their mutations in the inosine monophosphate dehydrogenase gene.

| Country | Organ transplant group<sup>a</sup> |  |  | Control group |  |  |
| --- | --- | --- | --- | --- | --- | --- |
|  | Total no. of isolates<sup>a</sup> | No. (%) of isolates on MPA | No. (%) of isolates with *impdh* mutations | Total no. of isolates | No. (%) of isolates on MPA | No. (%) of isolates with *impdh* mutations |
| USA | 9 | 8 (88·9) | 7<sup>b</sup> (77·8) | 6 | 0 (0) | 0 (0) |
| Denmark | 24 | 21 (87·5) | 17 (70·8) | 18 | 0 (0) | 3 (16·7) |
| Germany | 13 | 13 (100) | 13<sup>b</sup> (100) | 6 | 0 (0) | 2 (33·3) |
| Switzerland | 28 | 20<sup>c</sup> (71·4) | 25<sup>b</sup> (89·3) | 6 | 0 (0) | 2 (33·3) |
| Japan | 20 | 20 (100) | 19 (95·0) | 1 | 0 (0) | 0 (0) |
| China | 2 | 2 (100) | 2 (100) | 30 | 0 (0) | 2 (4·0) |
| *Total* | *96* | *84 (87·5)* | *83 (86·5)* | *67* | *0 (0)* | *9 (13·4)* |

<sup>a</sup>All involved in outbreaks except the two from China; all from renal transplants except for 11 from liver transplants in Denmark. <sup>b</sup>Including isolates containing multiple types of mutations at the same site or multiple sites (at codons 108, 261, 285, and 289), either as dominant or minor variants. <sup>c</sup>Excluding seven for which mycophenolic acid (MPA) usage was unknown (two from Denmark<sup>9</sup> and five from Switzerland<sup>15</sup>).

The SOT group consisted of 96 patients (Table 1): 94 patients involved in ten separate PCP outbreaks from the USA,^6^ Denmark,^9^ Germany,^10,11^ Switzerland,^7,10,15^ and Japan,^8,10,16^ and two from China who were not associated with an outbreak.^17,18^ Relevant clinical and epidemiological information about these outbreaks has been documented in the cited references. All were from kidney transplant recipients except for eight liver transplant recipients from Denmark. No patients received anti-*Pneumocystis* prophylaxis prior to PCP diagnosis. The vast majority of these patients (>89%) received mycophenolate mofetil (MMF, the prodrug of MPA) or MPA as part of the immunosuppressive regimen to prevent graft rejection at the time of PCP diagnosis.

The control group included 67 non-transplant patients with PCP. Thirty of them (17 from Denmark,^9^ six from Switzerland,^7,10,15^ six from Germany,^10,11^ and one from Japan^8,10,16^ were diagnosed during the same period at the same hospitals or cities, where outkreaks occurred. For 14 controls and 49 SOT recipients, multi-locus sequence typing (MLST) and/or restriction fragment length polymorphism (RFLP) analysis have been reported previously.^7,9–11^

### Identification of mutations in the *P. jirovecii impdh* gene

The *P. jirovecii impdh* gene was amplified by PCR as detailed in Supplementary Methods. All positive PCR products were initially sequenced by Sanger sequencing, with selected PCR products further sequenced by NGS. For eight previously sequenced isolates,^17–19^ NGS data were downloaded from public genome database (Supplementary Table S3).

### Multi-locus sequence typing (MLST) of *P. jirovecii* isolates

To determine if nonsynonymous mutations in the *P. jirovecii* impdh gene were associated with particular *P. jirovecii* strains, we performed MLST with the following genetic loci: nuclear internal transcribed spacer 1 (ITS1), 5.8S rRNA and ITS2 of the rRNA operon (ITSs), dihydropteroate synthase (*dhps*), superoxide dismutase (*sodA*), mitochondrial large subunit rRNA (mtLSU), cytochrome b (*cob*), and a highly polymorphic non-coding (PNC) region of the mitogenome (Supplementary Methods). For selected samples, the complete *P. jirovecii* mitogenome (mtDNA)^6,20^ was sequenced and highly discriminatory RFLP analysis^10,21^ was performed (Supplementary Methods).

### Structural modeling of wild-type and mutant IMPDH proteins

IMPDH protein sequences from different *Pneumocystis* species and other organisms were retrieved from the NCBI protein database or the UniProt database. The structure of wild-type *P. jirovecii* IMPDH was modeled using SWISS-MODEL, with the crystal structure of Chinese hamster IMPDH as the template (Supplementary Methods). Following optimization, the refined wild-type model was used as the template for modeling IMPDH proteins with either single mutations or combinations of two or more of the six common mutations (including H108Y, A261T, A261S, L285I, L285V and Q289L). Effects of mutations on protein stability were predicted using eight structure/sequence-based algorithms. Effects of mutations on IMPDH-MPA binding affinity were investigated by a series of computational approaches, including CB-DOCK2 molecular docking to predict optimal binding poses, CSM-lig and PRODIGY-LIG to estimate binding affinities, and PremPLI, and mCSM-lig to quantify mutation-induced changes in binding strength (Supplementary Methods). In addition, the MPA binding was assessed by superimposing the docking model onto the human IMPDH2 structure in complex with NAD^+^.

### Statistical analysis

Differences in the frequency of categorical variables were compared using Chi-square test or Fisher’s exact test as appropriate unless noted otherwise. A two-tailed p < 0·05 was considered as statistically significant. All analyses were carried out using SAS 9.4 (SAS Institute, Cary, NC, USA).

### Ethical considerations

The guidelines of the US Department of Health and Human Services and the NIH were followed in the conduct of the present study.

## Results

### Extensive inter- and intra-strain nucleotide polymorphisms in the *P. jirovecii impdh* gene

Initial Sanger Sequencing of either the full-length or the central region of the *impdh* gene in *P. jirovecii* isolates from 96 SOT patients and 67 controls identified single nucleotide polymorphisms (SNPs) at 16 positions, including 13 in exons and three in introns (Table 2). Non-synonymous changes (referred to as mutations hereafter) were present at seven positions. Subsequent NGS of *impdh* PCR products from 58 isolates (32 from SOT patients and 26 controls) identified additional SNPs as minor populations (with 1-49·9% of the total reads) from 25 patients, which included eight mutations at seven positions (Supplementary Table S4).

**Table 2:**
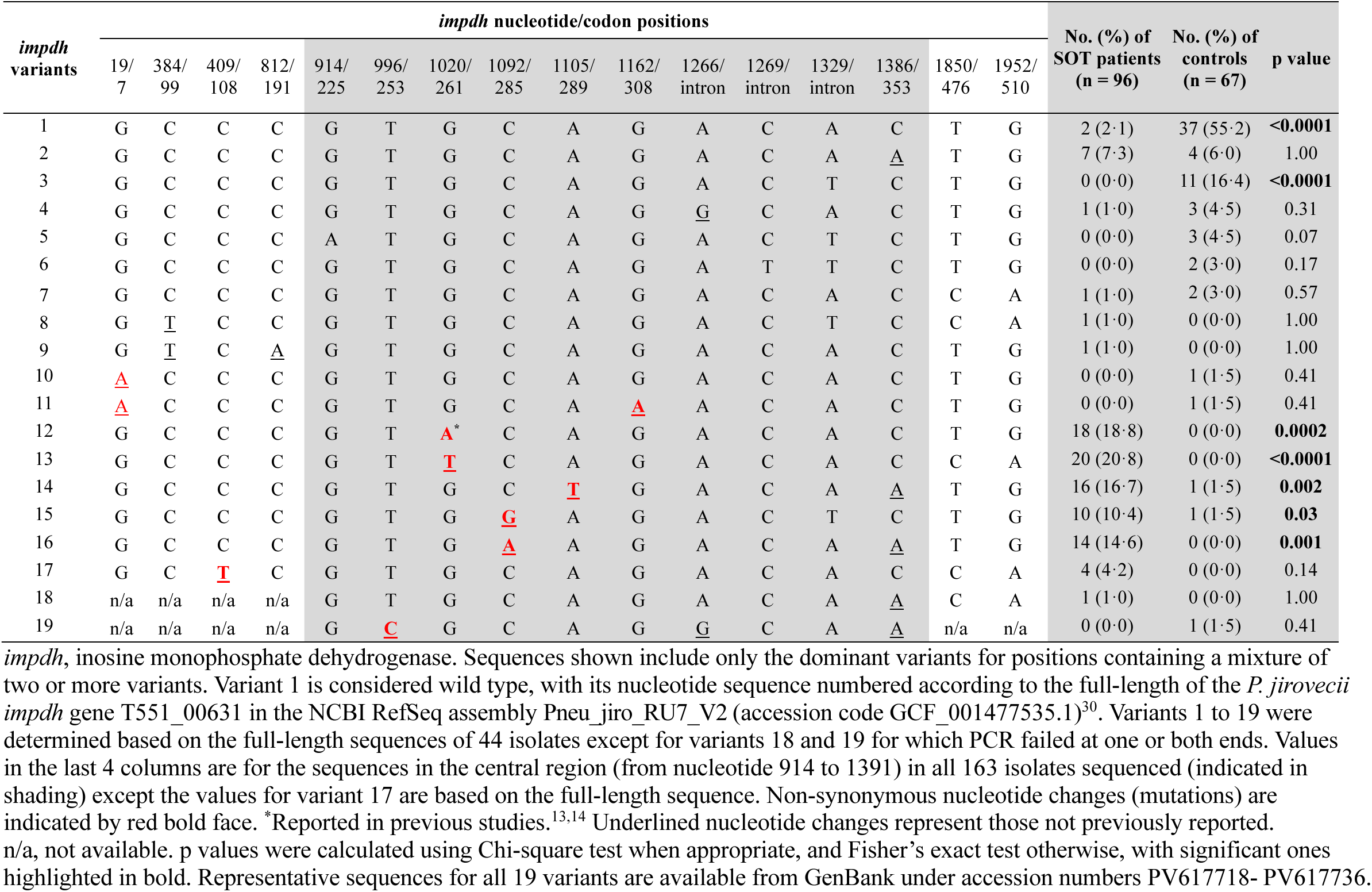
*P. jirovecii impdh* variants identified from solid organ transplant patients and non-transplant controls.

| *impdh* variants | *impdh* nucleotide/codon positions |  |  |  |  |  |  |  |  |  |  |  |  |  |  |  | No. (%) of SOT patients (n = 96) | No. (%) of controls (n = 67) | p value |
| --- | --- | --- | --- | --- | --- | --- | --- | --- | --- | --- | --- | --- | --- | --- | --- | --- | --- | --- | --- |
|  | 19/7 | 384/99 | 409/108 | 812/191 | 914/225 | 996/253 | 1020/261 | 1092/285 | 1105/289 | 1162/308 | 1266/intron | 1269/intron | 1329/intron | 1386/353 | 1850/476 | 1952/510 |  |  |  |
| 1 | G | C | C | C | G | T | G | C | A | G | A | C | A | C | T | G | 2 (2.1) | 37 (55.2) | **<0.0001** |
| 2 | G | C | C | C | G | T | G | C | A | G | A | C | A | <u>A</u> | T | G | 7 (7.3) | 4 (6.0) | 1.00 |
| 3 | G | C | C | C | G | T | G | C | A | G | A | C | T | C | T | G | 0 (0.0) | 11 (16.4) | **<0.0001** |
| 4 | G | C | C | C | G | T | G | C | A | G | <u>G</u> | C | A | C | T | G | 1 (1.0) | 3 (4.5) | 0.31 |
| 5 | G | C | C | C | A | T | G | C | A | G | A | C | T | C | T | G | 0 (0.0) | 3 (4.5) | 0.07 |
| 6 | G | C | C | C | G | T | G | C | A | G | A | T | T | C | T | G | 0 (0.0) | 2 (3.0) | 0.17 |
| 7 | G | C | C | C | G | T | G | C | A | G | A | C | A | C | C | A | 1 (1.0) | 2 (3.0) | 0.57 |
| 8 | G | <u>T</u> | C | C | G | T | G | C | A | G | A | C | T | C | C | A | 1 (1.0) | 0 (0.0) | 1.00 |
| 9 | G | <u>T</u> | C | <u>A</u> | G | T | G | C | A | G | A | C | A | C | T | G | 1 (1.0) | 0 (0.0) | 1.00 |
| 10 | <u>A</u> | C | C | C | G | T | G | C | A | G | A | C | A | C | T | G | 0 (0.0) | 1 (1.5) | 0.41 |
| 11 | <u>A</u> | C | C | C | G | T | G | C | A | <u>A</u> | A | C | A | C | T | G | 0 (0.0) | 1 (1.5) | 0.41 |
| 12 | G | C | C | C | G | T | **A\*** | C | A | G | A | C | A | C | T | G | 18 (18.8) | 0 (0.0) | **0.0002** |
| 13 | G | C | C | C | G | T | <u>T</u> | C | A | G | A | C | A | C | C | A | 20 (20.8) | 0 (0.0) | **<0.0001** |
| 14 | G | C | C | C | G | T | G | C | **T** | G | A | C | A | <u>A</u> | T | G | 16 (16.7) | 1 (1.5) | **0.002** |
| 15 | G | C | C | C | G | T | G | **G** | A | G | A | C | T | C | T | G | 10 (10.4) | 1 (1.5) | **0.03** |
| 16 | G | C | C | C | G | T | G | <u>A</u> | A | G | A | C | A | <u>A</u> | T | G | 14 (14.6) | 0 (0.0) | **0.001** |
| 17 | G | C | **T** | C | G | T | G | C | A | G | A | C | A | C | C | A | 4 (4.2) | 0 (0.0) | 0.14 |
| 18 | n/a | n/a | n/a | n/a | G | T | G | C | A | G | A | C | A | <u>A</u> | C | A | 1 (1.0) | 0 (0.0) | 1.00 |
| 19 | n/a | n/a | n/a | n/a | G | **C** | G | C | A | G | <u>G</u> | C | A | <u>A</u> | n/a | n/a | 0 (0.0) | 1 (1.5) | 0.41 |

*impdh*, inosine monophosphate dehydrogenase. Sequences shown include only the dominant variants for positions containing a mixture of two or more variants. Variant 1 is considered wild type, with its nucleotide sequence numbered according to the full-length of the *P. jirovecii* *impdh* gene T551\_00631 in the NCBI RefSeq assembly Pneu\_jiro\_RU7\_V2 (accession code GCF\_001477535.1)<sup>30</sup>. Variants 1 to 19 were determined based on the full-length sequences of 44 isolates except for variants 18 and 19 for which PCR failed at one or both ends. Values in the last 4 columns are for the sequences in the central region (from nucleotide 914 to 1391) in all 163 isolates sequenced (indicated in shading) except the values for variant 17 are based on the full-length sequence. Non-synonymous nucleotide changes (mutations) are indicated by red bold face. \*Reported in previous studies.<sup>13,14</sup> Underlined nucleotide changes represent those not previously reported. n/a, not available. p values were calculated using Chi-square test when appropriate, and Fisher's exact test otherwise, with significant ones highlighted in bold. Representative sequences for all 19 variants are available from GenBank under accession numbers PV617718- PV617736.

All previously reported *impdh* SNPs^13,14^ were identified in this study except for those at nucleotide positions 76, 1,737 and 1,754 (Table 2). Additionally, 12 novel SNPs, including nine mutations, were discovered. Combining the dominant SNPs at all 16 positions gave rise to 19 unique *impdh* variants. A total of ten mutations were identified at eight positions, including one detected exclusively as a minor variant in all nine samples sequenced (K225N, Supplementary Table S4). Six of these (H108Y, A261T, A261S, L285I, L285V, and Q289L) were detected at high frequencies (4-21% each) in SOT patients, appearing as dominant variants (with >50% NGS reads or highest peaks in Sanger sequencing). The remaining mutations were very infrequent (∼1% each in controls only) or only present as a minor variant (K225N) and were excluded from further analysis.

NGS identified two or more of the 6 common mutations coexisting within the same sample from 15 patients (11 SOTs and four controls); however, none of these isolates exhibited multiple mutations all present as dominant populations (Supplementary Table S4).

### Association of *P. jirovecii impdh* mutations with SOT and prior MPA exposure

In the Danish outbreak involving 24 SOT recipients and 17 controls, both mutations (L285V and Q289L), either individually or combined, were significantly more frequent in SOT recipients or those with MPA exposure than in controls or those without MPA exposure (p < 0·0001–0·01 in all comparisons, Table 3, Supplementary Table S6).

**Table 3:**
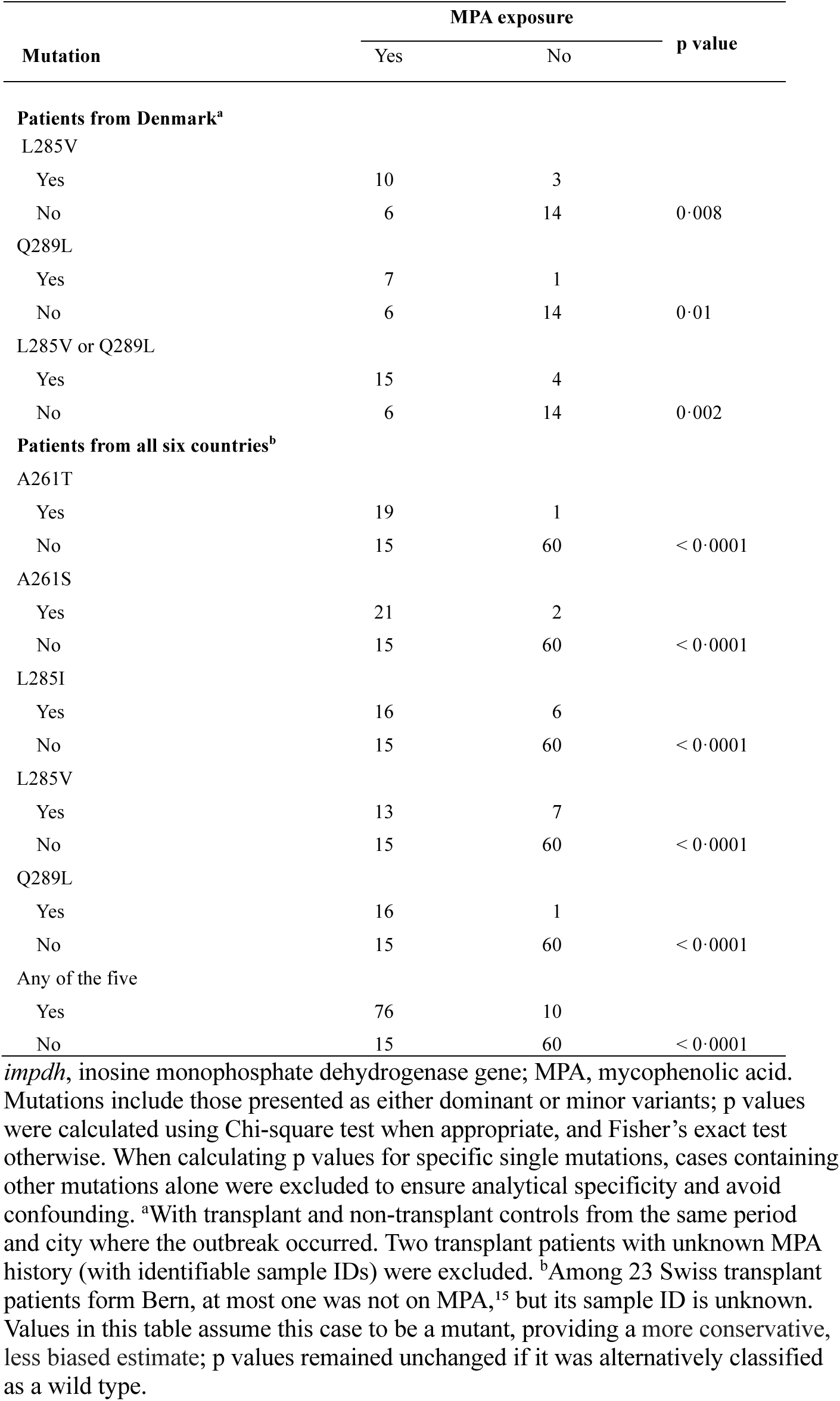
Correlation between *P. jirovecii impdh* mutations and MPA use.

When combining data from all six countries, the five common mutations (A261T, A261S, L285I, L285V, and Q289L), either individually or combined, were significantly more frequent in SOT recipients or those with MPA exposure than in controls or those without MPA exposure (p < 0·0001– 0·03 in all comparisons, Tables 2-3). The H108Y mutation did not show a significant difference in frequency between the SOT and control groups, likely due to the small sample size; its association with MPA exposure was not assessed due to the absence of relevant exposure information.^15^

### Association of *P. jirovecii impdh* mutations with specific strains

We performed extensive MLST typing at five genetic loci, integrated with full mtDNA and RFLP analysis in selected isolates. The combination of genotypes across all five loci or a subset including the two most polymorphic loci (ITSs and PNC) achieved perfect or near-perfect strain differentiation, with a discriminatory power of 99·89-100% (Supplementary Tables S7-S10). The high frequency of coinfection with multiple strains as revealed by MLST complicated the analysis of minor sequence populations (Supplementary Table S4). To overcome this difficulty, our genotype profiling focused on the mutations present as dominant variants, under the assumption that the dominant strain is likely responsible for the disease.

Our MLST analysis found that four of the six common mutations (except H108Y and L285V) were each associated with two to four strains (Table 4), which was further supported by full mtDNA and RFLP analysis in selected isolates (Supplementary Table S12-S13, Figure S1). There were a total of 11 strains associated with the six mutations. The previously reported mutation A261T^13^ was identified in 20 isolates in this study, with 19 linked to a single strain (designated EU8) and the remaining one linked to a different strain (designated CH55). Strain EU8 was identical to the strain with mutation L285I from one USA isolate. This was confirmed by MLST, full mtDNA, and RFLP analysis (Table 4, Supplementary Tables S12-S13). Similarly, the two isolates with different mutations (A261T and L285I) from two distant cities in China also appeared to be the same strain as supported by MLST and mtDNA analyses. These findings suggest the emergence of different mutations within the same strain.

**Table 4:** Genotype profiles of *P. jirovecii* isolates with inosine monophosphate dehydrogenase mutations.

| *impdh* mutations<sup>a</sup> | No. of isolates | Countries (no. of isolates) | *impdh* variant types | MLST allele profiles<sup>b</sup> | No. of isolates with mitogenome sequenced<sup>c</sup> | Strain designation |
| --- | --- | --- | --- | --- | --- | --- |
| A261T | 17 | Switzerland (16), USA (1) | 12 | 1-2-7-1-1 | 3 | EU8 |
|  | 1 | China (1) | 12 | 2-2-9-3-n | 1 | CH55 |
| A261S | 14 | Germany (12), Switzerland (2) | 13 | 1-2-1-18-53 | 3 | GS5 |
|  | 2 | Japan (2) | 13 | n-1-9-1-53 | 1 | JP35 |
|  | 1 | USA (1) | 13 | 1-2-8-3-53 | 1 | US6 |
| L285I | 8 | Japan (8) | 16 | 2-2-9-3-22 | 0 | JP18 |
|  | 4 | USA (4) | 16 | 2-1-9-3-22 | 4 | US1 |
|  | 1 | USA (1) | 16 | 1-2-7-1-1 | 1 | EU8 |
|  | 1 | China (1) | 16 | 2-2-9-3-6 | 1 | CH55 |
| L285V | 11 | Denmark (11) | 15 | 1-1-6-20-4 | 3 | DK2 |
| Q289L | 8 | Denmark (8) | 14 | 2-2-9-3-5 | 2 | DK3 |
|  | 9 | Japan (9) | 14 | 2-1-9-3-17 | 2 | JP38 |
| H108Y | 4 | Switzerland (4) | 17 | 2-1-1-32-17 | 0 | SW9 |

<sup>a</sup>Excluding mutations present as minor populations. All from transplant patients except for two controls from Denmark (D023 and D030). <sup>b</sup>Allele types at five genetic loci are detailed in Supplementary Tables S7-S10; one allele type for the Chinese isolate with A261T is not available (indicated by n). <sup>c</sup>The mitogenome sequences are identical between two or more isolates with identical multi-locus sequence type (MLST) profiles but different between isolates with different MLST profiles (Supplementary Table S12). Identical genotype profiles (including mitogenome) between isolates with different *impdh* mutations are highlighted with the same colors. Notably, the mitogenomes in two isolates with A261T from Switzerland (SB12) and the USA isolate (Y14) were identical to each other and to that of another USA isolate (Y16) with L285I.

Strain EU8 accounted for 21% (17/81) of all isolates carrying the six mutations, while all other strains accounted for 1-17%. Some patients were coinfected with two or more distinct strains, each harboring the same or different mutations, as exemplified in Supplementary Table S11. Given that the other four mutations were each linked to multiple strains, the observation that L285V and H108Y were each linked to a single strain may reflect limitations in sampling size, or geographic or temporal coverage.

### Geographic segregation and temporal shifts of *P. jirovecii impdh* mutations

The distributions of the six mutations differed significantly across six ountries (p < 0.0001, Supplementary Tables S15-S16, Figure 1). None were present in all six countries surveyed; however, three (A261T, A261S and L285I) were found across three continents (Europe, USA and Asia) while two (L285V and Q289L) were shared between Europe and Asia. H108Y was exclusively detected in Switzerland. Switzerland harbored five mutations while Japan had four. All other countries had no more than three. Considering the identity of the 11 strains with mutations (Table 4), strain EU8 was the only one shared across continents (Europe and North America) whereas all others were each confined to single countries except one strain (GS5), which was identified in Germany and Switzerland.

**Figure 1:**
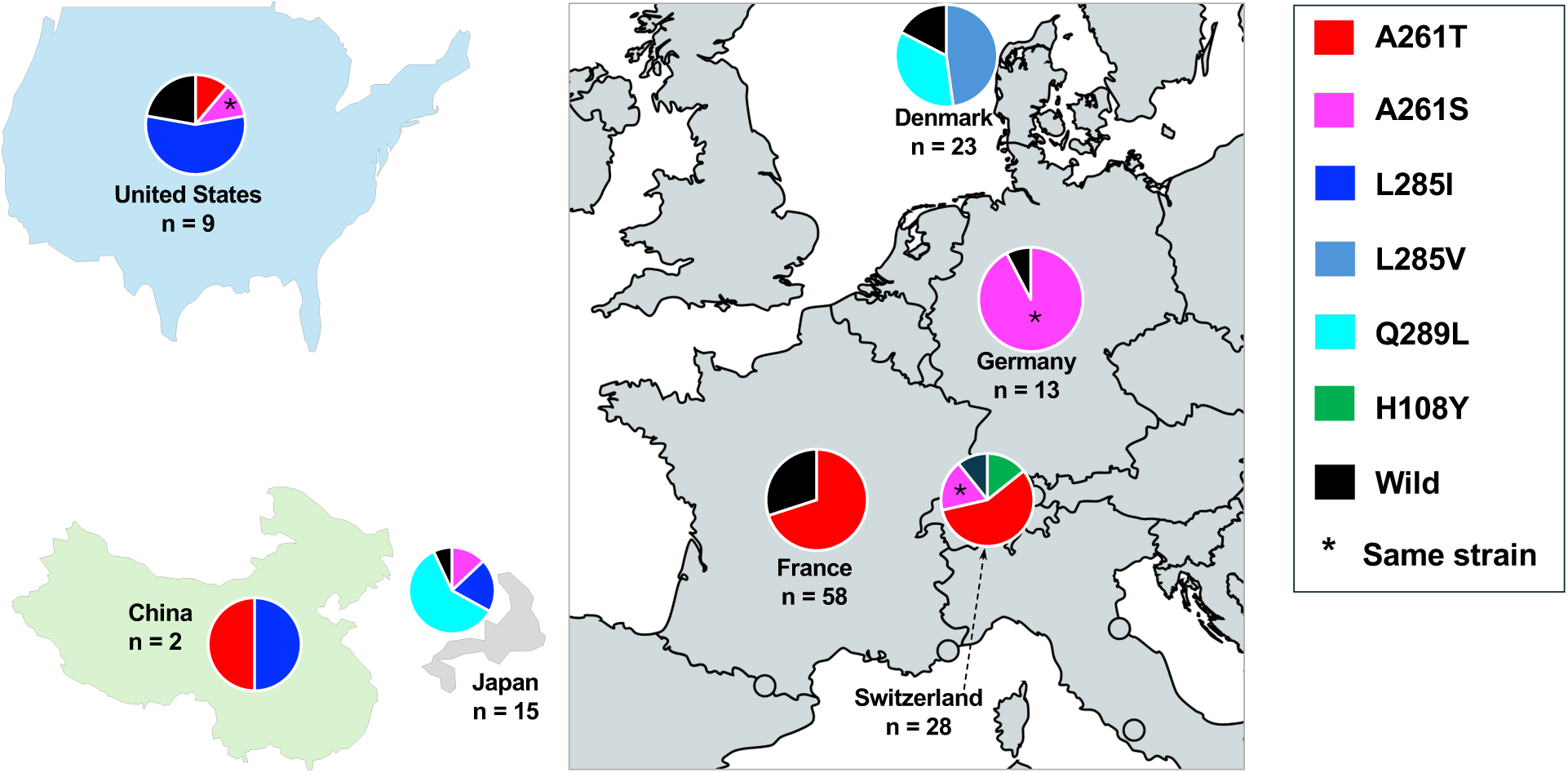
Distribution of P. jirovecii inosine monophosphate dehydrogenase (impdh) mutations in solid organ transplant recipients across different countries. Each pie shows the proportions of wild-type and mutant samples from transplant recipients (excluding mutations present as minor populations. See more detail in Supplementary Table S15). The number of cases from outbreaks are indicated, except for the cases in China, which were not associated with an outbreak. The 58 cases from France were reported at an international conference by Nevez et al. 2023. ^14^

In addition to geographic variation, there were apparent temporal shifts in mutations, either over different stages of an outbreak or between consecutive outbreaks in the same region or country (Figures 2-3, Supplementary Table S17). In a single outbreak in Denmark, two strains lacking *impdh* mutations were confined to the early phase, whereas two mutant strains (with L285V and Q289L respectively) emerged sympatrically during the late phase (Figure 2). In Japan, three separate outbreaks occurred in the same hospital between 2005 and 2014, each primarily associated with a unique strain with mutations L285I, A261S and Q289L, respectively (Figure 3). Similarly, three outbreaks in Switzerland (two in Bern and one in Zurich) from 2005 to 2012 exhibited a shift of mutations from H108Y to A261S, followed by A261T. Notably, the A261T strain in the 2007 outbreak in Zurich was shared with the 2006 outbreak in Munich, Germany, as previously reported.^10^

**Figure 2:**
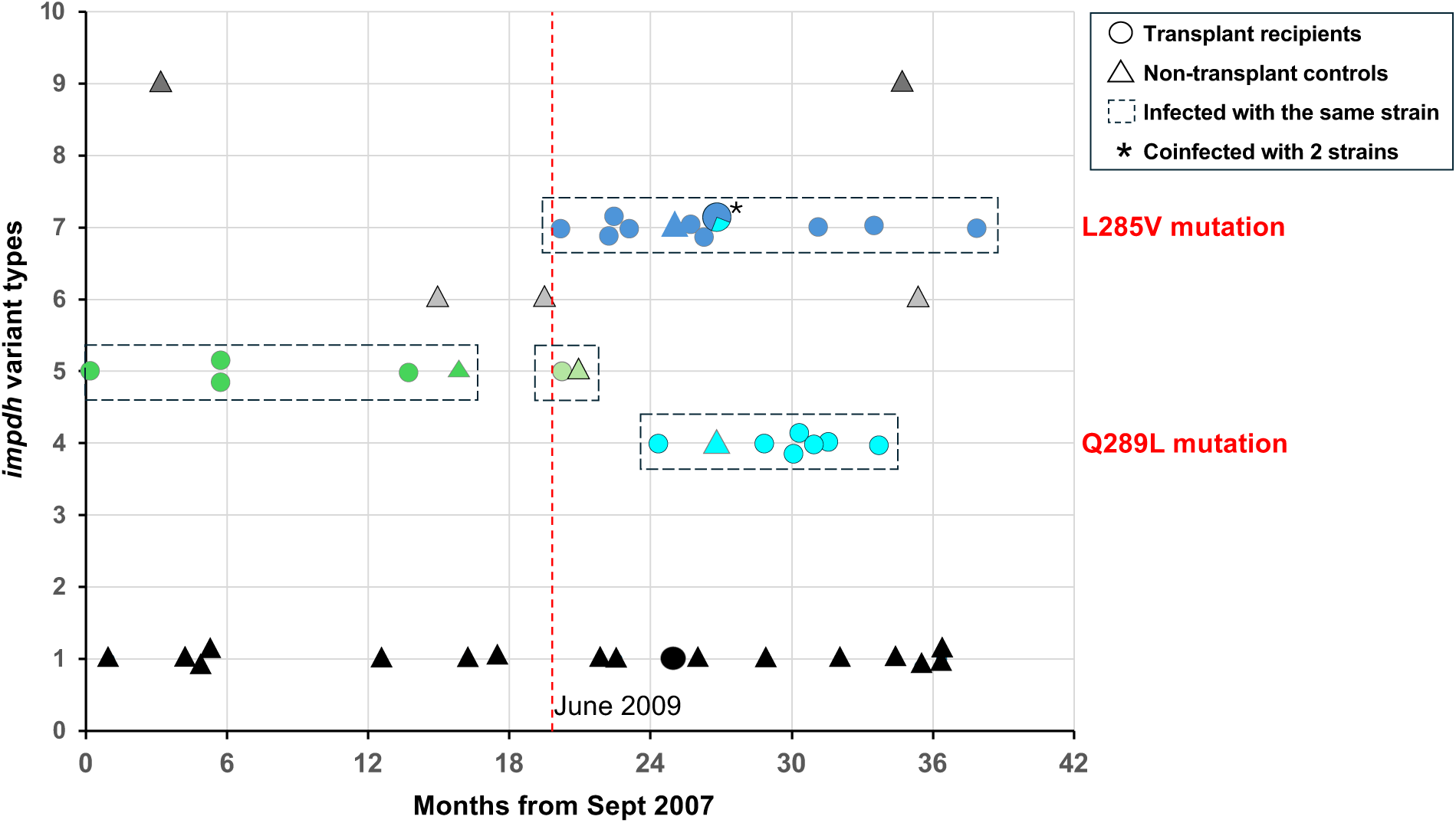
Shift of *P. jirovecii* strains from wild type to mutant during the outbreak among solid organ transplant recipients in Copenhagen, Denmark from 2007 to 2010. Each circle and triangle represent an individual solid organ transplant patient and non-transplant control, respectively. The numbers on the y-axis represent inosine monophosphate dehydrogenase (*impdh*) variant types as shown in Table 2; the same variants may be associated with different genotypes at other genetic loci as shown in Table 4. Isolates with the same genotypes (representing the same strains) are color-coded and boxed, including two strains with mutations L285V and Q289L. The patient (D024) marked by an asterisk is an example of patients coinfected with two mutant strains as detailed in Supplementary Table S11. Minor *impdh* variants in other isolates are omitted to enhance visualization. The vertical dotted line marks the onset of strains with *impdh* mutations.

**Figure 3:**
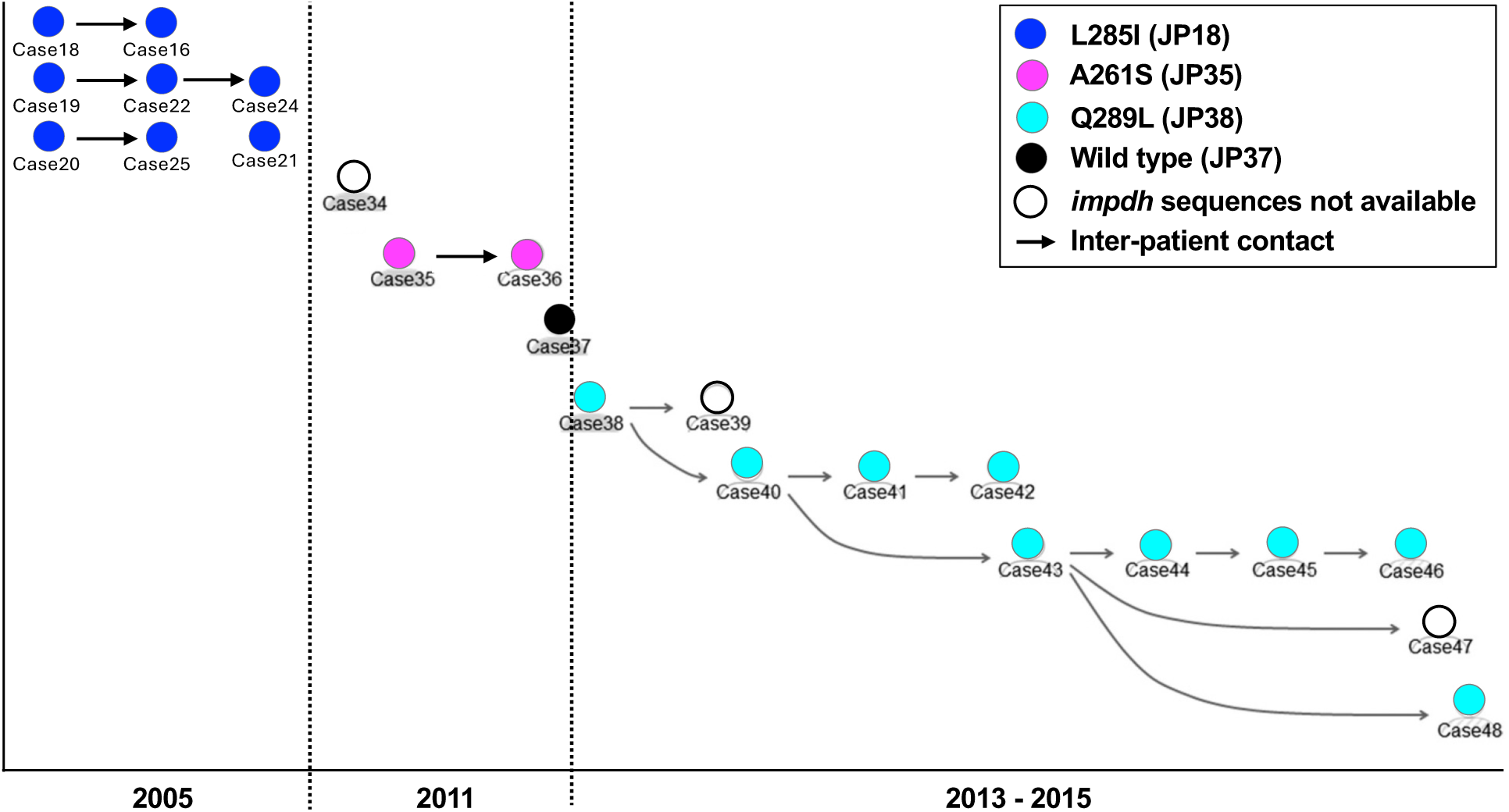
Shifts of *P. jirovecii* inosine monophosphate dehydrogenase (*impdh*) mutations during three consecutive outbreaks in renal transplant recipients in Nagoya, Japan from 2005 to 2015. This figure is adapted from reference^8^ by adding *impdh* mutation information. Each circle represents a renal transplant patient. Arrows indicate inter-person contacts. Wild-type and mutant *impdh* sequences are color-coded as shown in the top right corner, with strain IDs shown in parentheses (corresponding to those in Table 4 and Supplementary Table S17). Note that all cases with known contacts were infected with the same strains.

### Structural modeling of wild-type and mutant IMPDH proteins

Comparative analysis of IMPDH protein sequences revealed that all six common mutations were located within highly conserved regions of the enzyme catalytic domains shared across eukaryotic species, ranging from fungi to mammals (Supplementary Figure S2)). Mutations at these sites in several fungal species have been associated with resistance to MPA.^22,23^

To investigate the potential effects of the six common mutations on the protein structure and its binding affinity to MPA, we performed structural modeling analysis. Four mutations exhibited a destabilizing effect relative to the wild type, while the other two (H108Y and Q289L) had stabilizing effects, likely due to the tyrosine and leucine side chain forming more favorable hydrophobic interactions with Ala-261 and Met-337 compared to histidine and glutamine, respectively (Figure 4, Supplementary Figures S3-S4). Subsequent molecular docking analysis demonstrated that all other mutations, whether single or in combination (double and triple mutations), reduced binding affinity. H108Y and Q289L showed a binding score similar to that of the wild type. Further docking analysis revealed that mutated residues could distort the NAD^+^ binding site conformation, thus reducing E-XMP* accumulation and further impairing MPA binding. These observations, combined with binding affinity predictions and previous reports,^13,23,24^ sugges that these mutations contribute to MPA resistance.

**Figure 4:**
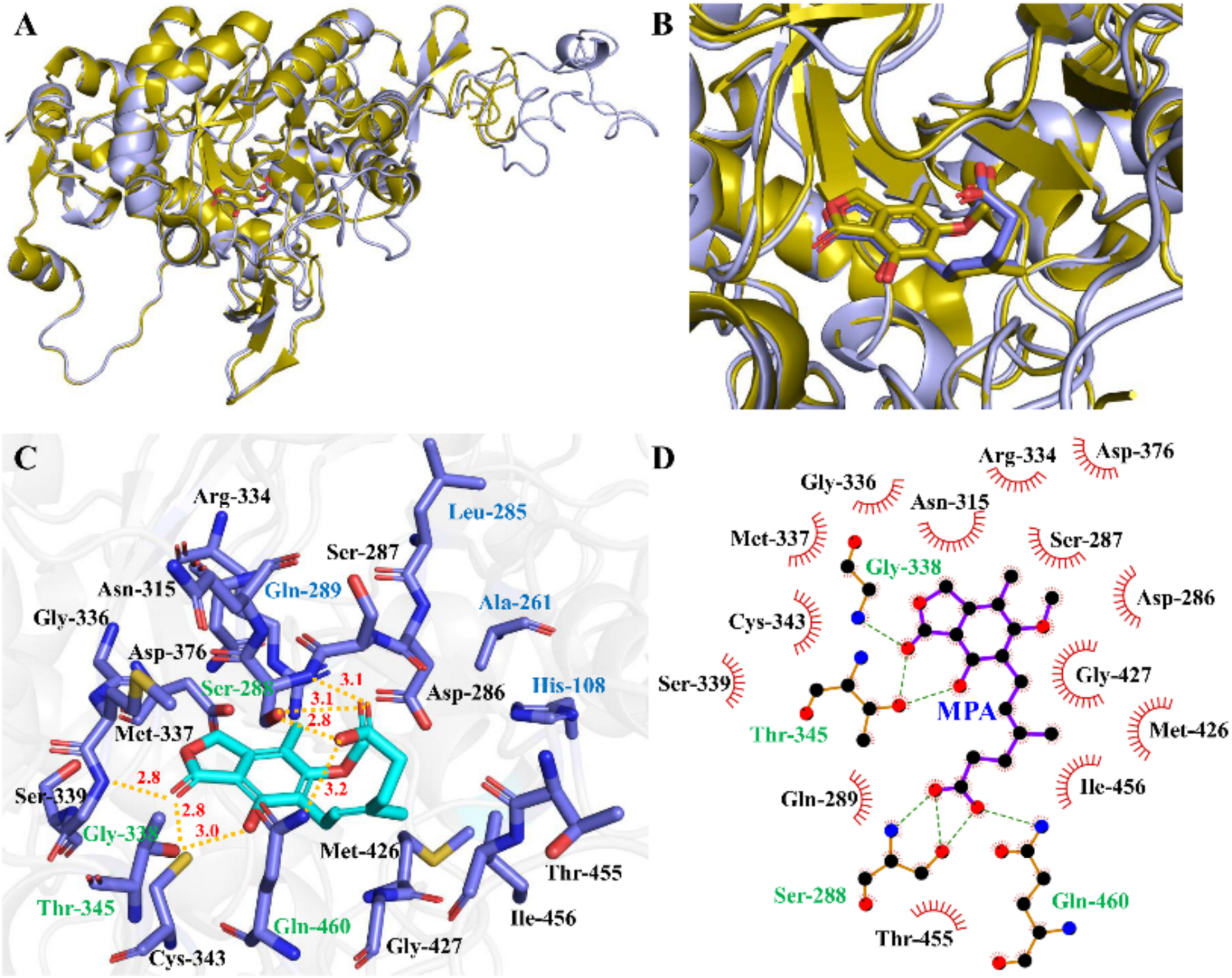
Structural modeling of inosine monophosphate dehydrogenase (IMPDH) and mycophenolic acid (MPA) binding in *P. jirovecii*. (**A**) Comparison of docking model of wild-type IMPDH and MPA with the crystal structure of 1JR1 (a reference structure of IMPDH from the Chinese hamster. Supplementary Methods). The docking model is shown in light blue while 1JR1 is shown in olive. The MPA binding site in the center is highlighted with a more detailed view in panel B. (**B**) Magnified view of the MPA binding site within IMPDH. The docked MPA (planar aromatic structure in light blue by element) exhibits a high degree of structural congruence with the experimental MPA (olive by element) from 1JR1. (**C**) Detailed binding site analysis where MPA is colored in cyan and residues in slate. Key residues within 5 Å around MPA are shown, with Ala-261 and Leu-285 positioned at a distance exceeding 5 Å. Hydrogen bonds are represented in dashed yellow lines and distances are labeled in red. The residues forming H-bonds with MPA are colored in green. The four residues that exhibit mutations in clinical samples are labeled in blue. (**D**) 2D representation of the binding site from panel C, generated by LigPlot+, where hydrogen bonds are shown in dashed green lines. Due to the critical role of the four amino acids (H108, A261, L285, and Q289) in stabilizing the conformations of adjacent residues, some of which (e.g., Ser-288) form strong interactions with MPA, the mutations observed in clinical samples (H108Y, A261T, A261S, L285I, L285V, Q289L) likely affect the MPA binding by altering the conformations of neighboring residues.

## Discussion

In this international, multicenter study, we analyzed *P. jirovecii* samples from 163 immunocompromised individuals, including 96 SOT recipients and 67 non-transplant controls, across six countries spanning Europe, North America, and Asia. The majority of the SOT recipients received MPA therapy and were involved in nine separate outbreaks between 2005 and 2020. Our analysis revealed six common mutations in the *P. jirovecii impdh* gene, including the previously reported A261T. Five mutations were strongly associated with SOT status and MPA exposure. All mutations displayed marked geographic and temporal variations during outbreaks. More importantly, our study demonstrated that four of the six mutations were each linked to multiple distinct strains. Furthermore, structural modeling predicted that four mutations likely reduce protein stability and binding affinity to MPA.

IMPDH plays a critical role in purine biosynthesis and is highly conserved across multiple species spanning from fungi to mammals; MPA inhibits IMPDH activity, preferentially inhibiting proliferation of lymphocytes and resulting in immunosuppression though it also has anti-*Pneumocystis* activity as discussed below. In this study, we identified six mutations in the *P. jirovecii impdh* gene, with an overall prevalence of 87% (83/96) among SOT patients across six countries, significantly higher than the 69% (40/58) reported in French patients with the A261T mutation (p = 0·012. Supplementary text).^14^ This suggests that these mutations confer a growth advantage in this setting, likely by conferring resistance to MPA, as supported by the following observations. First, in vitro studies using short-term cultures of *P. carinii* from rats and recombinant *P. carinii* IMPDH protein demonstrated significant inhibition of IMPDH activity by MPA,^25^ providing direct evidence of the drug’s effect on the *Pneumocystis* enzyme. This effect is further supported by an in vivo study in a rat model where MPA protected the animal against *Pneumocystis* infection.^26^ Second, multiple clinical trials evaluating the efficacy of the MPA prodrug MMF in preventing graft rejection in SOT patients soon after its approval showed a lower incidence of PCP in MMF-treated patients compared to untreated controls,^27^ supporting an inhibitory effect of MMF on *P. jirovecii,* assuming no *impdh* mutations or resistance existed at that time (MPA was not approved for use in SOT until 1995). Third, the strong association between these mutations and prior MPA exposure, as observed in this and previous studies, suggests that the mutations were selected under MPA exposure. Fourth, our structure modeling suggests that four of the six *impdh* mutations in *P. jirovecii* lead to reduced protein stability and binding affinity to MPA (Figure 4, Supplementary Tables S18-S19 and Figures S3-S4). Finally, studies in other organisms, such as *Candida albicans*, *Saccharomyces cerevisiae* and *Penicillium brevicompactum*, have shown that the mutation homologous to A261T in *P. jirovecii* can be induced *in vitro* by exposure to MPA and confers resistance to MPA (Supplementary Figure S2);^22^ a mutation analogous to Q289L in *P. jirovecii* has been identified in MPA-resistant mutants (Q277R) of human IMPDH2.^28^ Together, these observations strongly suggest that MPA can exert selective pressure on *P. jirovecii*, potentially leading to the emergence of drug resistance. These findings represent a rare instance of a human cell-targeted drug exerting unanticipated cross-species effects on a microbial pathogen, highlighting the broader implications of human-fungal cell similarity and the intricate interplay between pharmacological interventions, pathogen evolution, and strain selection.

While further studies are required to confirm the MPA resistance in *P. jirovecii*, our findings support the hypothesis of selection and maintenance of MPA-resistant *P. jirovecii* strains in SOT patients, which confers a selective growth advantage in immunosuppressed patients receiving MPA, thus contributing to transmission and outbreaks within this population, as summarized in Figure 5. This study provided compelling evidence for the emergence of *impdh* mutations in approximately a dozen *P. jirovecii* strains, with variable prevalence in different countries (Table 4 and Supplementary Tables S15-S16). The identification of identical strains between SOT patients with contact strongly supports *de novo* infection rather than reactivation of a latent infection and suggests a high transmissibility of the mutant strain (Figure 3). The high prevalence of coinfection with multiple strains (Supplementary Table S4) suggests a strong selective pressure by MPA or a potential accelerated evolution of mutations and drug resistance in *P. jirovecii* as has been demonstrated in other pathogens.^29^ The observed switch from wild type to mutant strains as well as between strains with different mutations during outbreaks (Figures 2 and 3, Supplementary Table S17) suggests the possibility of widespread dissemination of these mutations globally. It remains to be determined what factors trigger the mutation switch (Supplementary text) and whether the emergence of multiple mutations within the same strain or across different strains in the same patient, together with geographic and temporal variations, could confer any advantage in maintaining or enhancing MPA resistance and thereby facilitate transmission and outbreaks.

**Figure 5:**
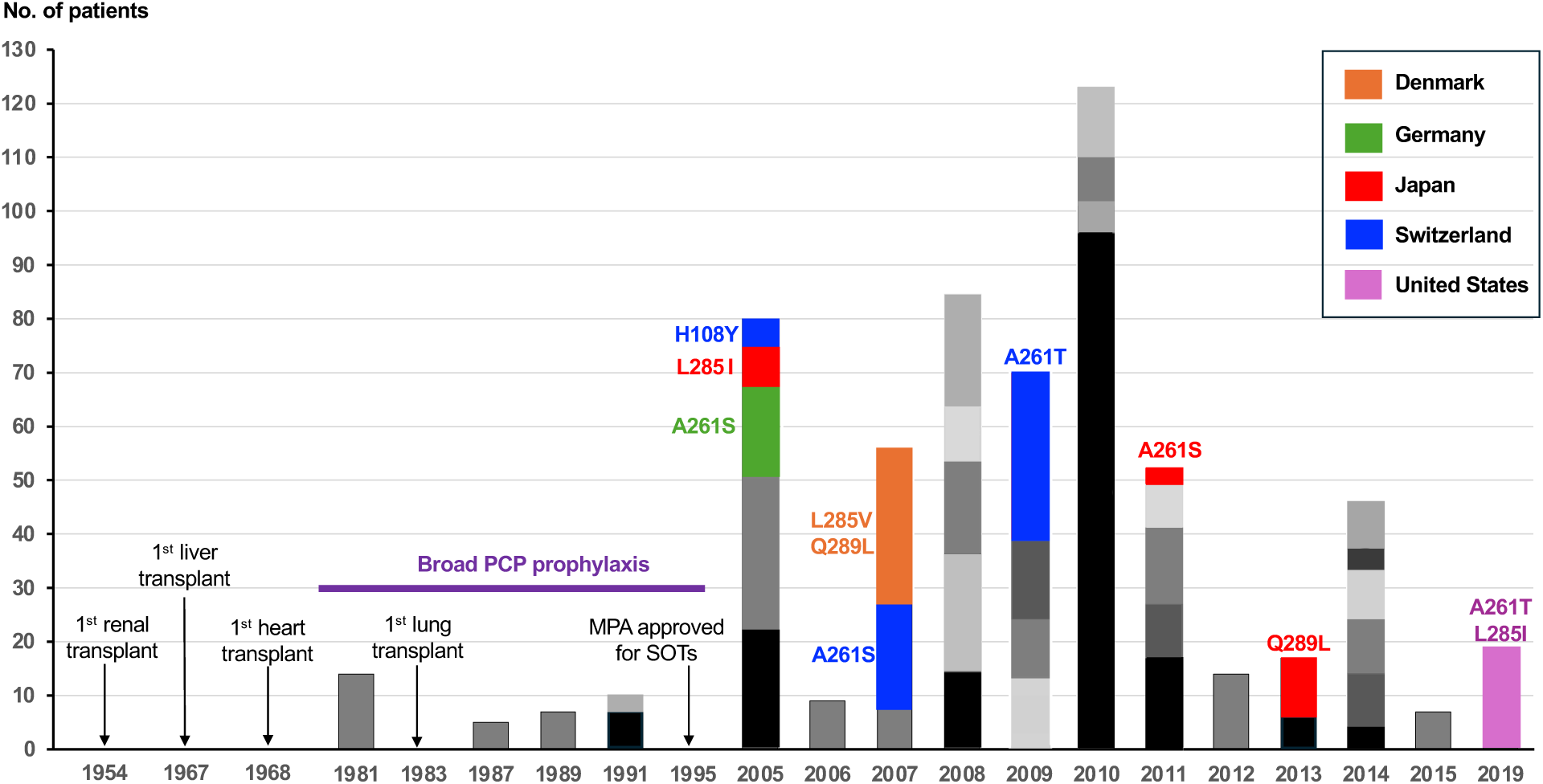
Timeline of *Pneumocystis* pneumonia (PCP) outbreaks in solid organ transplant recipients and emergence of *P. jirovecii* inosine monophosphate dehydrogenase (*impdh*) mutations. Outbreak cases shown are according to all reports from 1981 to 2020 (detailed in Supplementary Table S20). Each vertical bar denotes a single outbreak, with the year of onset indicated on the x-axis and the number of PCP patients corresponding to the scale on the y-axis. Stacked bars represent multiple separate outbreaks that started in the same year. Colored bars denote the nine separate outbreaks from five countries (color coded as shown in the upper right corner) investigated in this study, with associated *impdh* mutations indicated above or to the left of the bars (with the same color coding). Note there were three independent outbreaks with distinct *impdh* mutations occurred in Switzerland as well as Japan (detailed in the Supplementary Table S17). Major milestones for organ transplantation are also marked. Before 2005, only sporadic, small-scale outbreaks occurred due to widespread prophylaxis. Some transplant clinics discontinued prophylaxis when cases declined; however, no major outbreaks occurred before 2005, possibly owing to the adoption of MPA for graft rejection, which also has potent anti-*Pneumocystis* activity. Over time, MPA likely exerted selective pressure, leading to *P. jirovecii impdh* mutations that potentially confer resistance, diminishing MPA’s anti-*Pneumocystis* effects. Therefore, mutant strains could easily spread among transplant recipients on MPA, while wild-type strains remained suppressed. Multiple mutations arose independently in diverse strains with the potential to switch between and within strains, likely enhancing resistance and facilitating transmission and outbreak development.

Understanding the role of MPA in selecting specific *P. jirovecii* strains in SOT patients has important clinical implications. Exposure to MPA should be recognized as a significant risk factor for the development of PCP involving MPA-resistant strains in SOT patients and others receiving this drug. To mitigate this risk, prolonged PCP chemoprophylaxis and patient isolation precautions (especially at transplant clinics) may be critical for MPA-treated patients. Avoiding cohorting of SOT recipients in the same room can potentially minimize transmission. Healthcare facilities, particularly SOT clinics, might consider establishing molecular surveillance to monitor for *P. jirovecii impdh* mutant strains in high-risk populations. Furthermore, given its potential to increase PCP risk and promote *impdh* mutation development, the prophylactic use of MPA in SOT for organ rejection prevention might merit cautious reconsideration; exploration of alternative strategies may be warranted.

The major strength of this study lies in the use of a large number of samples with a broad global geographic coverage and a relatively long temporal span, enabling the identification of both known and novel mutations as well as their variable geographic distribution patterns. This is notably the largest molecular study of *Pneumocystis* outbreaks conducted to date. Additionally, the integration of comprehensive genotyping with NGS, full mitogenome and RFLP analyses enhanced strain identification and differentiation, particularly by confirming of linkage of individual mutations to multiple strains. Furthermore, comparative IMPDH protein motif analysis and structural modeling offered valuable insights into the potential molecular mechanisms underlying MPA resistance. However, the study also has notable limitations, including the lack of detailed epidemiological and clinical background data for many patients, relatively small sample sizes and lack of appropriately matched controls from several outbreaks, limited genomic data on *P. jirovecii* mutants, and no testing of the functional effects of the mutations. These limitations constrain the ability to draw precise and generalized inferences, underscoring the critical need for more rigorous data collection in future research endeavors.

In conclusion, we identified six common mutations in the *P. jirovecii impdh* gene, strongly associated with MPA exposure and predicted to alter protein stability and its binding affinity for MPA. These mutations, arising independently in multiple *P. jirovecii* strains, are spreading globally with variable geographic distributions and temporal shifts, potentially representing a major mechanism of resistance contributing to strain selection, transmission, and outbreaks in SOT settings. These findings have significant implications for clinical management and prevention strategies of PCP. Moreover, our study illustrates a rare example of how antimicrobial resistance may evolve through unanticipated pathways, extending beyond typical antimicrobial use, warranting heightened awareness and strategic consideration in clinical practice.

## Data Availability

All sequences generated in this study were deposited into the NCBI database, with accession numbers listed in Table 4 and Supplementary Tables S7-S8 and S12.

## Contributors

LM and JAK conceived the project and designed the methodology. MMA, AC, XD, TF, SG, NG, JHL, CH, LH (Huang), RK, NJM, SO, LAP, LP, AAR, AS, YW, and HY collected the samples and patient clinical data. LM, XD, LFW, BS and GH performed PCR and DNA sequencing. MS performed the RFLP analysis. WC, MH and LH (Hedstrom) carried out computational structure modeling and drug resistance prediction. LM and JS conducted statistical analysis. JAK and TI provided resources and supervision. LM drafted the manuscript, with all authors critically contributing to the review and editing process and providing final approval for publication. All authors had full access to all the data in the study and had final responsibility for the decision to submit for publication.

## Data sharing statement

All novel sequences generated in this study were deposited into the NCBI nucleotide database, with accession numbers listed in Table 4 and Supplementary Tables S7-S8 and S12).

## Ethical considerations

The National Institutes of Health Institutional Review Board and the National Institute of Allergy and Infectious Diseases Institutional Review Board gave ethical approval for the *Pneumocystis* protocol. The protocol is registered with Clinicaltrials.gov, ID# NCT00342589. The Office of Human Subjects Research Protections, Office of Intramural Research, Office of the Director, National Institutes of Health waived ethical approval for the samples provided from outside institutions. The guidelines of the US Department of Health and Human Services and the NIH were followed in the conduct of the present study. The authors confirm that the sample IDs and sample collection dates were not known to anyone outside the research team involved in this study.

## Acknowledgements

The views, information or content, and conclusions presented do not necessarily represent the official position or policy of, nor should any official endorsement be inferred on the part of, the Clinical Center, the National Institutes of Health, or the Department of Health and Human Services.

