## Supplemental Methods, Tables and Figures for "Unanticipated Global Emergence of Multiple *Pneumocystis jirovecii* Mutants Selected by Mycophenolic Acid Driving Increasing Outbreaks in Solid Organ Transplant Recipients"

### 1. Genetic loci and reference sequences used in this study

**1.1.** *P. jirovecii* inosine monophosphate dehydrogenase (*impdh*) gene

A previous study of the full-length sequence (2,070 bp) of the *P. jirovecii impdh* gene in 26 patients (including 10 solid organ transplant recipients and 16 non-transplant controls) identified single nucleotide polymorphisms (SNPs) at nine positions, including nonsynonymous (missense) mutations at three codons, A26S, A261T and G439S.[1] A261T was identified in all ten transplant recipients but was absent in the controls while the other two mutations were identified only in controls (one and three patients, respectively) and located at positions that varied among different *Pneumocystis* species and among other species.[1] Given that A261T was located in a highly conserved position across all species and within the catalytic domain of the IMPDH enzyme and that homologous mutations in other organisms have been linked to MPA resistance,[2] our study focused primarily on the central region of 682 bp encompassing this position, which could be efficiently amplified by PCR.

Following the identification of multiple mutations in addition to those reported previously,[1] we sequenced the remaining portions of both ends of this gene in a subset of 44 samples representating different groups of isolates with diverse *impdh* mutations and/or genotypes.

**1.2.** Genetic markers for *P. jirovecii* multi-locus sequencing typing (MLST)

Based on their discriminatory power, PCR accessibility, compatibility with existing data, and coverage of both nuclear and mitochondrial genomes (mtDNA), we selected the following genetic markers: nuclear internal transcribed spacer 1 (ITS1), 5.8S rRNA and internal transcribed spacer 2 (ITS2) of the rRNA operon (collectively referred to as ITS1-5.8S-ITS2 or ITSs), dihydropteroate synthase (*dhps*), superoxide dismutase (*sodA*), mtDNA large subunit rRNA (mtLSU),[3] and an ~1.0-kb highly polymorphic non-coding (PNC) region of mtDNA.[4,5] *dhps* was included for an additional reason - its potential implication in drug resistance.[6-8]

**1.3**. Genetic target for restriction fragment length polymorphism (RFLP)

The RFLP system targets an ~1.4-kb fragment of the *P. jirovecii* major surface glycoprotein gene family (*msg*-A1), which consists of ~86 members per genome. [9,10] This system has demonstrated exceptional discriminatory power in strain typing.[4,11-13] However, this method has two major drawbacks: it requires at least 1,000 *msg*-A1 gene copies per PCR reaction to yield reliable results, and it cannot differentiate coinfections involving multiple high-abundance *P. jirovecii* strains.

**1.4.** *P. jirovecii* full mtDNA sequencing

For selected *P. jirovecii* isolates, full mtDNAs were sequenced to strengthen strain typing.

**1.5.** Reference sequences used in this study

All reference sequences used are from the *P. jirovecii* RU7 nuclear and mitochondrial genomes, which represent the most complete genome assemblies available for *P. jirovecii* to date.[5,9] The nucleotide positions for *impdh* mutations and SNPs are relative to the full-length sequence of the gene.

| Genetic loci | Supercontig ID | NCBI Reference Sequence ID | Nucleotide positions | Notes |
|---|---|---|---|---|
| *impdh* | Supercont1.3 | NW_017264777.1 | 46207-48276 | Locus tag T551_00631 |
| *dhps* | Supercont1.9 | NW_017264783.1 | 368362-367529 | Part of the *fas* gene, locus tag T551_02230 |
| *sodA* | Supercont1.12 | NW_017264786.1 | 135572-136576 | Locus tag T551_02671 |
| ITS1-5.8S-ITS2 (or ITSs) | Supercont1.2 | NW_017264776.1 | 611059-611546 | Between locus tags T551_05004 (18S rRNA) and T551_05005 (26S rRNA) |
| mtLSU | Mitogenome | NC_020331.1 | 12373-15076 | Gene ID *rnl* or locus tag H866_mgr02 |
| PNC | Mitogenome | NC_020331.1 | 931-1946 | Non-coding region |

### 2. PCR amplification

The sequences of all oligonucleotide primers used are detailed in Supplementary Table S1. The main PCR conditions are summarized in Supplementary Table S2.

Universal PCR precautions were taken to avoid potential contamination, including the use of separate workplaces for pre-PCR and post-PCR steps, aliquoting of primers and other reagents for one-time use, setting-up of reaction mixtures in a dedicated hood, and inclusion of negative controls in each experiment.[14]

To amplify targets < 2kb, the primary PCR was conducted using the 2X LiTaq Plus PCR Master Mix (LifeSct LLC, Rockville, MD, USA) and the thermocycling program as follows: 95 °C for 2 min; 10 cycles of 94°C for 30s, 65°C for 60 s with 1.5°C decease per cycle, and 72 °C for 2 min; 30 cycles of 94°C for 30s, 5°C for 30s, and 72°C for 1.5 min; and a final extension at 72 °C for 5 min. PCR products were separated by agarose gel electrophoresis using the 1% E-Gel with SYBR Safe DNA stain (Thermo Fisher Scientific, Waltham, MA, USA). For samples showing weak or no bands in agarose gel electrophoresis, nested- or semi-nested-PCR was performed using the 2X LiTaq Plus PCR Master Mix and the thermocycling program as follows: 95 °C for 2 min; 30 cycles of 94 °C for 30s, 50 °C for 30s, and 72 °C for 1.5 min; and a final extension at 72 °C for 5 min.

To amplify overlapping fragments (2.3-5.4 kb) for mtDNA, PCR was conducted using the LongAmp Hot Start Taq 2X Master Mix (New England Biolabs, Ipswich, MA, USA) and the thermocycling program as follows: 95 °C for 2 min; 10 cycles of 94 °C for 30s, 55°C for 60 s with 1.0 °C decrease per cycle, and 65 °C for 7 min; 30 cycles of 94 °C for 30s, 45 °C for 30s, and 65 °C for 7 min; and a final extension at 65 °C for 10 min. To amplify the mtDNA non-coding region (>85% AT content), an additional 200 µM of dATP and dTTP was added and the extension temperature was reduced to 55°C.

### 3. Sanger sequencing and next-generation sequencing (NGS)

All positive PCR products were purified and sent to Poochon Scientific (Frederick, Maryland, USA) for direct Sanger sequencing from both directions. Selected samples, including those showing mixed sequence populations in initial Sanger sequencing as well as representatives from clusters showing different genotype profiles (described below in section 5) and all PCR products for the full mtDNA sequencing, were further sequenced using commercial NGS services by LifeSct LLC (Rockville, Maryland, USA) and AmpSeq (Gaithersburg, Maryland, USA). For NGS, all PCR products amplified from the same sample were pooled together as one amplicon mixture. NGS was conducted using the Illumina NovaSeq system (Illumina Inc., San Diego, CA, USA) and Element AVITI system (Element Biosciences, San Diego, CA, USA). All libraries were sequenced with 150-base paired-end reads, with a minimum of 1Gb data per library.

**4. Characterization of *P. jirovecii impdh* mutations.**

**4.1**. Methods of *impdh* sequence analysis

For Sanger sequencing data, chromatogram trace files (abi format) were aligned to the *P. jirovecii impdh* reference sequence described above using the Sequencher (5.4.6., Gene Codes Corp, Ann Arbor, Michigan, USA). All SNPs relative to the reference were recorded. For samples showing mixed sequences, minor SNPs were identified as peaks clearly exceeding the baseline in chromatograms from both directions. For samples that were also sequenced by NGS, SNP calls were determined as described below.

For NGS data, all raw reads were aligned to the *P. jirovecii impdh* reference sequence using SeqMan NGen (version 18.0.3, DNASTAR, Madison, WI, USA) with the Variant Analysis/Resequecing workflow under default settings except that the minimum match threshold the Assembly Options was set to 93%.

Since amplicon NGS data are prone to sequence errors resulting from both PCR amplification and the sequencing process, we tested the baseline error rates in order to accurately identify low-frequency mutations in samples containing mixed sequence populations. This test involved NGS sequencing of *P. jirovecii impdh* PCR products from non-transplant controls with no MPA exposure and not involved in any outbreak, including a pooled mixture of PCR products from 7 isolates, with a total of 1,326,873 reads (150 bp each), and individual PCR products from 12 isolates, with 3,949-386,123 reads each (150-bp read length). By Sanger sequencing, all these PCR products showed homogeneous sequences with no indication of mixed populations at any nucleotide positions showing non-synonymous mutations from SOT patients. Based on NGS reads, the frequency of minor mutations in the pooled PCR products varied from 0·04% to 0·35%; the highest frequency of minor mutations in individual PCR products varied from 0·18% to 0·7%. In addition, the highest frequency of minor SNPs in a position within an intron (nucleotide 1282, unlikely affected by drug pressure) was 0·6% among all PCR products sequenced by NGS, including those from SOT patients with MPA exposure. Based on these observations, a threshold of 1% was set for calling a minor mutation. The same threshold was applied for SNP calling in MLST analysis as described below. SNPs present in >50% of reads were defined as dominant SNPs (or mutations or alleles) in all samples containing mixed sequence populations.

In samples containing a mixture of three or more *impdh* alleles, it is unfeasible to reliably assemble the short NGS reads into the full length of the *impdh* amplicon for the minor alleles. To overcome this limitation, we analyzed the NGS reads spanning the three nucleotide positions with the 5 common mutations (at nucleotide 1020/codon 261, nucleotide 1092/codon

285, and nucleotide 1105/codon 289) in 58 samples. NGS raw reads spanning all these three nucleotide positions were retrieved using SeqMan NGen by mapping against the *P. jirovecii imdph* reference sequence. Retrieved reads were assembled using Sequencher with a minimum match percentage of 99%. Each contig was checked manually to remove reads containing any ambiguous bases at or near the three mutation sites, and the number of reads for each contig was counted. This allowed the identification of double mutations present as minor populations exemplified in the Supplementary Table S5.

4.2. Prevalence of *impdh* mutations compared to that of *dhps* mutations

The results of this study support previous reports demonstrating the emergence of mutations in the *P. jirovecii* i*mpdh* gene,[1,15] and provide new insights into the mutation profiles and prevalence dynamics. We identified a total of ten mutations in 85 of 163 (52%) *P. jirovecii* isolates, including 84 of 96 (88%) from SOTs and 9 of 67 (13%) from controls (including mutations present as minor populations). There were six common mutations identified in 84 of 96 (88%) isolates from SOTs, and 8 of 67 (12%) from controls, indicating a high prevalence of mutations in *P. jirovecii* within this patient population. Only one of these mutations (A261T) has been reported previously, primarily among French patients,[1,15] with a prevalence of 69% (40/58) in SOT recipients and complete absence in controls. In our study, this mutation showed a prevalence of 20% (19/96) in SOTs and was absent in controls. The five novel mutations identified in our study each had a prevalence of 4-21% in SOTs and up to 2% in controls.

Combining the data in this and previous studies,[1,15] the overall prevalence of all six mutaitons was 80% (123/154) in SOTs (mostly with MPA exposure) and 7% (9/126) in controls, which appear to be similar to the prevalence of *dhps* mutations (at codons 55 and 57), 62-84% in patients with sulfa exposure[6,16,17] and 3-10% in those without.[17,18] This may reflect a comparable level of selection pressure on *P. jirovecii* exerted by sulfa drugs and MPA, consistent with the widespread use of both drugs. It is noteworthy MPA has been prescribed not to prevent or treat PCP, but rather for its immunosuppressive effects in preventing graft rejection. The linkage of four of the six common mutations with multiple strains suggests that each of them did not originate from a single *P. jirovecii* strain but instead emerged independently across multiple distinct strains, also similar to the emergence of the *dhps* mutations.[19]

4.3. Implications of geographic and temporal variations in impdh mutations.

The use of a larger number of samples with a broader geographic and temporal representation in this study than in previous studies enabled the identification of not only novel mutations but also pronounced geographic and temporal variation in their distribution. What factors contributed to this variation cannot be determined in this study. It could be related to the limited number of samples in certain regions, variations in MPA dosage and ingredients, or differences in drug pharmacokinetics and pharmacodynamics between different patient populations.[20] Alternatively, it may simply represent a stochastic event occurring in a local strain. It is particularly interesting to observe the temporal shift from a wild-type strain to a mixture of two strains with mutations Q289L and L285V in the outbreak in Denmark (Figure 2). This shift may suggest a selection process of the mutant strains. However, the selection spread is not expected to be fast given the inherently slow replication and growth rates of *Pneumocystis* organisms and the relatively small population of SOT recipients within the larger MPA-unexposed population. Based on the timeline of mutations observed in outbreaks in Denmark and Japan, this selection process may take approximately one to two years.

It is also interesting to observe the broad distribution of the most common mutation A261T across Europe, North America and Asia (Figure 1, Table 4). We provided strong evidence of the involvement of the same strain carrying this mutation in separate outbreaks across Europe (Switzerland) and North America (USA), as supported by MLST, full mtDNA data and RFLP analyses (Table 4, Supplementary Table S12). In addition, this strain shared an identical MLST profile and mitogenome with another strain carrying mutation L285I in the same outbreak in USA, suggesting that these two strains may have originated from the same strain but subsequently evolved different mutations over time. This possibility may also exist for the strain found in the two Chinese patients with mutations A261T and L285I.

Of note, 12 (12%) of the 98 isolates from SOT patient showed no *impdh* mutations but were associated with 6 distinct *impdh* variants due to nonsynonymous changes (Supplementary Table S14). They were involved in outbreaks from three countries (Denmark, Switzerland, and USA), with at least seven cases receiving MPA treatment. Each represented a unique strain except for five Danish isolates, which belonged to the same strain based on MLST, mtDNA and RFLP analyses (Supplementary Tables S12-S13), and one additional Danish isolate (D002) which was identical to the strain in a Danish control (D053).

The presence of multiple distinct *impdh* variants with no mutations indicates genetic heterogeneity, suggesting that nonsynonymous changes may contribute to *P. jirovecii* diversity in SOT patients. The distribution across multiple countries suggests independent spread or regional adaptation. The occurrence of unique strains, except for the clonal cluster of Danish isolates, indicates both clonal expansion and independent emergence. The detection of the same strain in an SOT recipient (D002) and a control patient (D053) during the Danish outbreak suggests person-to-person transmission.

**5. Determination of *P. jirovecii* MLST allele profiles**

SNPs at the five genetic markers were determined based on Sanger sequencing in combination with NGS for selected PCR products using the same methods described above for the *impdh* sequence analysis.

**5.1**. Genotypes at *dhps*, *sodA* and mtLSU

Three loci, including *dhps*, *sodA* and mtLSU, each contained only two to five polymorphic sites, with their nucleotide positions numbered relative to the full-length gene sequences of respective references listed above in Section 1.5. The position numbering for *dhps* (including positions 163, 169 and 513) is consistent with all previous reports [6,21]. For *sodA*, the positions 250 and 355 described in this study correspond to 110 and 215 in previous reports.[3] For mtLSU, the positions 843 and 1006 described in this study correspond to 85 and 248 in previous reports;[3] positions 1171 and 1182-1188 described in this study are not available from any previous genotyping studies. For a small number of samples from Japan (n = 9), Germany (n = 7), and Denmark (n = 9), *dhps* or mtLSU sequences were obtained from previous studies[12,22,23] but shorter than the amplicons in this study; their genotypes included in this study were limited to the positions determined in previous studies, with any undetermined positions marked as not available (n/a) in Table 4 and Supplementary Table S12-S14.

**5.2**. Genotypes at ITSs and PNC

Both ITSs and PNC showed extensive SNPs; unique sequences were assigned allele or genotype numbers as previously described by Xue *et al.*[24] and Azar *et al.*,[4] respectively. Genotypes identical to previously reported ones (1 to 62 for ITSs and 1 to 22 for PNC) were assigned the same numbers, while new genotypes were numbered sequentially, following the existing series (64 to 70 for ITSs and 23 to 67 for PNC, Supplementary Table S8). For ITSs genotypes, variation in the length of seven homopolymeric T or A tracts was omitted because it has been well documented that such variation can occur as an artifact during PCR and sequencing.[24,25] The PNC genotypes obtained in this study spanned the full length, or nearly the full length, of the amplicons, which replaced the previous short versions when the overlapping regions were identical. For samples containing mixed sequence populations, the dominant genotypes were determined from either the direct readout of Sanger sequencing or consensus sequences of aligned NGS reads; the dominance was verified by manual inspection of the chromatogram of Sanger sequencing or by comparing NGS read counts at polymorphic sites. Meanwhile, this process also enabled the identification of minor genotypes in samples containing only two sequence populations differing by with only a few SNPs.

For samples containing a mixture of three or more sequence populations, it was not feasible to reliably assemble the 150-base short NGS reads into the full-length sequence of the ITSs and PNC amplicons (623-1,028 bp). Therefore, we chose to focus on shorter, highly polymorphic regions of ~200-300 bp to estimate the number of unique sequence populations representing different *P. jirovecii* strains in selected samples. NGS raw reads for these regions were retrieved using SeqMan NGen by mapping against the dominant sequence as determined by Sanger sequencing under low stringent conditions (with a minimum match percentage of 73%). Retrieved reads were assembled using Sequencher with a minimum match percentage of 99% and a minimum overlap length of 90 bp. Each contig was checked manually to remove reads containing any mismatch at the positions defining a genotype, or without an overlap with the main reads for at least 90 bp. A contig containing at least 10 reads was used to define a genotype. All contigs meeting these requirements were aligned using MacVector 18.6.1 (MacVector Inc., Cary, NC, USA) to compare sequence divergence. The results of conifection in 58 isolates (35%) are summarized in Supplementary Table S4.

**5.3**. Discriminatory index of MLST

To determine the discriminatory power of MLST markers, we selected 40 *P. jirovecii* isolates from non-transplant controls with no known contact with infected transplant patients. For isolates contained mixtures of two or more genotypes, only the dominant genotype was considered. Discriminatory index (DI) was calculated using the method by Hunter *et al.*[26] via the webserver at http://insilico.ehu.eus/mini_tools/discriminatory_power/index.php. The results are provided in Supplementary Tables S9-S10. For individual markers, PNC showed the greatest discriminatory power (DI = 0·9956), followed by ITSs (DI = 0·9502). Combining these two or combining either one with all three others could provide sufficient discriminatory power (DI = 0·9989-1·0000), comparable to that achieved with combining all 5 markers (DI = 1·0000). For this reason, and owing to the limited availability of DNA samples, not all five markers were examined for every other sample. Nevertheless, all outbreak samples were genotyped at a minimum of three loci, resulting in an DI of > 0·99, thereby enabling confident determination of strain associations.

**5.4**. Allele assignment and strain definition

Like the genotypes at ITSs and PNC, each unique sequence at *dhps*, *sodA* and mtLSU was also assigned an allele number as detailed in Supplementary Table S7. The combination of allele numbers across all loci defines the isolate's allelic profile. For isolates containing multiple alleles at any loci, only the dominant alleles were used to define the allelic profile. Isolates with identical allelic profiles were considered the same strain (Table 4).

However, since not all 5 genetic markers were sequenced in every isolates as described above, any isoaltes showing different alleles at PNC and ITS or at either of them plus all three other loci were considered different strains while any samples showing identical alleles at these loci were considered the same strain. For example, the two isolates from China (Pj55 and S24) showed identical allelels at all loci except for the ITSs, which could not be determined due to insufficient NGS reads and unavailability of samples for testing in this study; these two isolates were considered the same strain (Table 4).

Notably, *dhps* mutations at codons 55/57 (including allelles 4-6, potentially associated with sulfa drug resistance[6,16,17]) were detected in 6 of 52 (12%) control patients tested but were absent in 85 SOT patients. It is unknown if these 6 patients had prior exposure to sulfa drugs while almost all SOT patients were not on PCP prophylaxis at the time of PCP diagnosis.

**5.5.** Coinfection with multiple *P. jirovecii* variants

*impdh* showed substantial variations not only between different isolates but also within individual isolates. Among the 165 *P. jirovecii* isolates examined, 38 (23%) contained a mixture of two or more SNPs (including the wild-type nucleotide) at one or more of the 13 positions with SNPs. The frequency of mixed SNPs (at any of the 13 positions) was significantly lower in SOT recipients than in controls (18/98 or 18% *vs* 23/67 or 34%, p = 0·02). However, when restricted to codons 261, 285 and 289 (the sites of the five most common mutations), the ferequency of mixed SNPs did not differ significantly between SOT recipients and controls (12/98 or 12% vs 7/67 or 10%, p = 0·83). The frequency of coinfection with multiple strains as determined by MLST (at any of the five loci) was significantly lower in SOT recipients than in controls (28/98 or 29% vs 32/67 or 48%, p = 0·01). These findings indicate that overall genetic diversity of *P. jirovecii*, including mixed SNPs at *impdh* and coinfection with multiple strains, is reduced in SOT recipients compared with controls. This pattern may reflect selective pressure from MPA, which could favor the outgrowth of dominant strains while limiting within-host diversity and suppressing minority strains. However, the lack of significant difference in the frequency of mixed SNPs at *impdh* codons 261, 285, and 289 implies that such selective effects may not uniformly impact all mutation sites.

### 6. Sequencing of *P. jirovecii* mtDNA

mtDNA reads were retrieved from NGS data by mapping against the reference mtDNA of *P. jirovecii* RU7 using SeqMan NGen under default conditions except for reducing the minimum match percentage to 70%. Retrieved reads were *de novo* assembled using SeqMan NGen under default conditions except for increasing the minimum match percentage to 97%. The resulting contigs were further assembled using Sequencher. Gaps were filled by multiple rounds of alignment of merged contigs to NGS raw reads. The final assembly was re-aligned to NGS raw reads to check for any potential assembly errors as well as SNPs. mtDNA annotation was performed using the MFannot tool with genetic code 4 (http://megasun.bch.umontreal.ca/cgi-bin/mfannot/mfannotInterface.pl). Transfer RNAs were further evaluated using tRNAscan-SE (128). All annotated genes were reviewed and compared with the homologs in the reference mtDNA of *P. jirovecii*.

A total of 40 *P. jirovecii* mtDNA sequences were included for comparative analysis, including 28 from SOT patients and 13 from non-transplant controls (Supplementary Table S12). Only three of them (from SOT patients in USA, Y1, Y6 anfd Y13) were partial (11-23 kb), whereas all others were complete (33-35 kb). Two of them were reconstructed in one isolate from a Danish SOT patient (D024). The mtDNAs from all non-transplant controls were markedly different from one another and from those of the SOT patients. In contrast, mtDNAs from SOT patients were essentially identical among isolates sharing the same MLST allele profiles, with only minor variations in the copy number of a few tandem repeats (mainly homopolymers) in some cases (Supplementary Table S12).

### 7. RFLP analysis

Previous studies have employed RFLP analysis on samples from PCP outbreaks in the USA (New Haven),[4] Denmark (Copenhagen),[12] Japan (Nagoya), Germany (Munich), and Switzerland (Zurich).[11,13] In this study, RFLP was conducted only on samples from the outbreak in Bern, Switzerland, following previously described methods. [11,13] Briefly, *msg*-A1 gene copy number in the DNA samples was quantified by a realtime quantitative PCR, and samples with a minimum of 1000 *msg*-A1 gene copies per PCR reaction were selected for RFLP analysis. PCR amplification was performed using a seminested PCR protocol with primers GK472, GK452, and GK195, as described. [11] PCR products were purified and then treated with restriction enzymes Dra1 or Hpy188I. Digested fragments were separated using 1.2% agarose gels. The results are shown in Supplementary Figure S1.

We correlated the RFLP patterns obtained in this study and previous studies with *impdh* mutations and MLST profiles as summarized in Supplementary Table S13. From all these studies, all samples with an identical RFLP pattern also showed an identical MLST profile, including samples between separate outbreaks in Zurich, Switzerland and Munich, Germany. Whereas, all samples with an identical MLST profile also showed an identical RFLP pattern except for one outbreak sample from Japan (J8) and another outbreak sample from the USA (Y16). In J8, the mtLSU genotype was not available but the genotypes at all the other 4 loci were identical to those of other three isolates from the same outbreak (J1, J2 and J5). Thus, these 4 isolates were considered the same strain due to the high discriminatory power of the four loci sequenced (DI = 1·0000. Supplementary Tables S9-S10). Sample Y16 contained a mixture of 7 strains, with the dominant strain showing an identical MLST profile as well as an identical mtDNA sequence to the strain found in the other three transplant patients from the same outbreak (Y1, Y3 and Y13); the difference in the RFLP pattern could be attributed to coinfection with additional strains possessing distinct *msg*-A1 repertoires.

Within each outbreak analyzed by RFLP, multiple *impdh* mutations were identified in the USA and Denmark while a single *impdh* mutation was observed in Switzerland (including two separate outbreaks in Bern and one outbreak in Zurich) and Germany; each mutation was linked to a specific RFLP pattern except for in the USA, Denmark and Japan, where the mutation L285I was linked to two distinct RFLP patterns. Of note, the outbreaks in Zurich, Switzerland and Munich, Germany shared the mutation A261S as well as an identical RFLP pattern (Supplementary Table S13).

### 8. Structural modeling of *P. jirovecii* IMPDH

**8.1.** Structure modeling of wild-type IMPDH and its mutants

IMPDH protein sequences from different *Pneumocystis* species and other organisms were retrieved from the NCBI protein database or the UniProt database (Supplementary Figure S2), and aligned using MacVector 18.6.1 (MacVector Inc., Cary, NC, USA). The structure of wild-type IMPDH for *P. jirovecii* strain RU7 (UniProt ID: A0A0W4ZU90) was modeled using SWISS-MODEL.[27] The structure with PDB ID of 1JR1[28] was used as the template for modeling due to its high homology with *P. jirovecii* IMPDH and inclusion of MPA as the ligand in the PDB structure. It is noteworthy that 1JR1 is from Chinese hamster IMPDH, which is similar with human type II IMPDH with only six amino acid differences and both have similar enzymatic characteristics. [28] After generating the initial wild-type homology model, YASARA[29] was employed to perform relatively large-scale optimization for improving the model. The YASARA-optimized model was then used as the template by SWISS-MODEL for modeling several IMPDH single, double, and triple mutants, including H108Y, A261T, A261S, L285I, L285V, Q289L, A261T/L285I, A261S/L285I, A261S/L285V, A261T/L285I/Q289L and A261T/L285V/Q289L. For comparison, the wild-type model was also predicted based on the YASARA-optimized template. To check the modeled structure, the wild-type IMPDH structure was aligned to chain A of 1JR1. As shown in Supplementary Figure S3, both structures superimpose together well with RMSD of 0·418 Å measured in open-source PyMOL (https://github.com/schrodinger/pymol-open-source).

**8.2.** Predicted mutation effects on protein stability

To investigate the mutation effects on protein stability, multiple structure/sequence-based algorithms, including DDMut,[30] DynaMut2,[31] DDGun,[32] Eris,[33] DUET,[34], MUpro,[35] mCSM,[36] and iStable [37] were used to perform the predictions. The first four methods were considered as the main tools because all of them were able to to perform predictions for either single, double, or triple mutations by using the wild-type IMPDH protein structure as the only input, where the last four methods were used to predict the single mutation effects on protein stability. As shown in Supplementary Table S18, the consensus (i.e., majority vote among all methods) was used as the final predictions. By comparison of the wild-type protein, it is evident that all mutations present the destabilizing effect except that from H108Y and Q289L. For the double mutations (i.e., A261T/L285I, A261S/L285I and A261S/L285V), the overlapping effect was also investigated using last four methods by setting A261T or A261S as the control structure, where the additional L285I or L285V was predicted by all last four methods to exhibit the destabilizing effect on protein stability. Of note, the predicted results for double mutations from DUET, MUpro, mCSM and iStable cannot be directly compared with those from the first four methods for double and triple mutations due to the differences in initial input structures. One possible reason that H1088Y and Q289L increase the protein stability is that the side chain of tyrosine and leucine could form more favorable hydrophobic interactions with nearby residues (e.g., Ala-261 and Met-337, respectively, Figure 4).

**8.3.** Predicted mutation effects on IMPDH-MPA binding affinity

To investigate the mutation effects on IMPDH-MPA binding affinity, the CB-DOCK2 molecular docking method [38] was used to predict the reasonable binding mode by considering all 12 modeled protein structures including wild-type and single, double, and triple mutants as the corresponding receptors and the MPA structure obtained from the RCSB PDB database[39] as the ligand. The final binding models were obtained by using the template-based method with the optimal CB-DOCK2 docking scores shown in Supplementary Table S19. For comparison of binding site with the experimental structure, the docking model of wild-type IMPDH and MPA was aligned to 1JR1 as shown in Figures 4A and 4B. The figures show that the docked MPA superimposes well with MPA from 1JR1, indicating the binding sites of MPA are similar for both proteins (RMSD = 0·26 Å of the two MPA molecules). This is supported by the strong conservation of the MPA binding site between fungal and human IMPDH enzymes.[40] Two other methods (i.e., CSM-lig[41] and PRODIGY-LIG[42]) were also used to predict the binding affinity of wild-type and mutants with MPA (Supplementary Table S19). In addition, PremPLI[43] and mCSM-lig,[44] two structure-guided in silico approaches, were used to quantify the effects of single mutations on affinities of IMPDH and mutants with MPA. Most single, double, and triple mutations cause slightly decreased binding affinity as predicted by CB-DOCK2, except for H108Y and Q289L, which exhibit the same binding score as wild-type. CSM-lig and PRODIGY-LIG predict wild-type and its mutants to have comparable binding affinity. PremPLI quantifies all single mutations to decrease the binding affinity, while mCSM-lig shows similar trend but for Q289L shows increased affinity between MPA and this mutant.

In summary, all mutations were predicted to decrease either the protein stability or ligand binding affinity except the Q289L mutant, which showed increased protein stability and ligand binding affinity by mCSM-lig, possibly due to the major hydrophobic interactions around this mutant (Figure 4C). The detailed binding model between wild-type IMPDH and MPA is shown in Figure 4C, where key residues within 5 Å around MPA are shown and additional two residues (i.e., Ala-261 and Leu-285, more than 5 Å with MPA) are also labeled in Figure 4C. Figure 4D was generated by LigPlot+,[45] showing 2D MPA and IMPDH interaction diagram.

To further investigate the mutation effects on MPA binding, the docking model was also aligned to the E•IMP•$NAD^+$ complex of human IMPDH2 (PDB ID: 6U8N)[46] as shown in Supplementary Figure S4A. It is evident that MPA binds to the site of nicotinamide portion from $NAD^+$, which is consistent with the previous study.[40] As reported, the MPA binding affinity depends on both the accumulation of intermediate E-XMP* and the structure of the MPA/$NAD^+$ binding site.[47] Thus, to investigate the residue positions of six mutations (H108Y, A261T, A261S, L285I, L285V and Q289L) in *P. jirovecii* IMPDH, we explored the key residues around $NAD^+$ with 5 Å based on the aligned structures. As shown in Supplementary Figure S4B, several representative key residues including His-108, Thr-264, Arg-265, Pro-266, Asp-286, Ser-287, Ser-288, Gln-289 and Tyr-294 are highlighted in our docking model. These residues are conserved, with the exception of Arg-265, Pro-266 and Tyr-294, which are His, Glu, and Phe, respectively, in human IMPDH2. The corresponding residues in 6U8N are His-93, Thr-252, His-253, Glu-254, Asp-274, Ser-275, Ser-276, Gln-277 and Phe-282, respectively.

In the current study, the observed missense mutation residues with higher frequencies are Ala-261, Leu-285 and Gln-289. Since Ala-261 resides at the end of a beta strand near to the Thr-264, Arg-265, Pro-266 loop, A261T or A261S mutations may affect the loop conformation. Interestingly, Leu-285 also resides at the end of a beta strand/beginning of a loop segment, including Asp-286, Ser-287, Ser-288 and Gln-289. Thus, the L285I or L285V mutations may change the loop conformation via a side chain steric interaction. Gln-289 residue interacts directly with $NAD^+$. Therefore, the Q289L mutation undoubtedly plays a major role in the $NAD^+$ binding. In addition, this switch of a polar to non-polar residues could favor hydrophobic interactions with other residues. In fact, this residue is also involved the MPA binding as shown in Figure 4.

In summary, the mutated residues could change the conformation of the $NAD^+$ binding site, directly affecting the binding of MPA and also likely perturbing the accumulation of E-XMP*.[47] By combination of the binding affinity prediction mentioned above and other reports,[1,40,47] these mutations likely cause MPA resistance.

### 9. Sequence data availability

All nucleotide sequences for new *P. jirovecii impdh* alleles or genotypes at five genetic markers as well as mitogenome sequences have been deposited into the NCBI database, with accession numbers provided in Table 4 and Supplementary Tables S7-S8 and S12. Sequences identical to existing ones at the NCBI database were assigned the same accession numbers.

**Supplementary Table S1: PCR primers used in this study**

| Genetic targets | Primer IDs | Nucleotide sequences (5' to 3') | References |
|---|---|---|---|
| ***Mitochondrial genome**** | | | |
| *cob* | cob.r8 | GGATTAGCCATRATRTAATTATCRCTATG | Ma *et al.*[14] |
| *cox2* | JM1r | GATGGTGCCAGTCCTGTTTT | This study |
| *cox2* | JM2f | GTCCGATCGAATCCTCAAAA | This study |
| *cox3* | cox3.f2 | AGAGAATGATGAGCATAAGT | This study |
| *cox3* | JM3r | TGCCTTGTCACCTGCTGTAG | This study |
| mtLSU | LSU.f1 | CTCATGTCAGCATTCTCTCTTTA | Ma *et al.*[14] |
| mtLSU | Pu3Y.f | TACAAATCGRACTAGGATAT | Ma *et al.*[14] |
| mtLSU | LSU.r1 | TAGGATATAGCTGGTTTTCTGCGA | Ma *et al.*[14] |
| mtLSU | mit.f6 | CTCACGGTACTCTTCACTA | This study |
| mtLSU | rnl.f1 | TWAACCCAACTCACG | This study |
| mtLSU | rnl.r2 | CACCTCGATGTCGACTCA | This study |
| mtLSU | rnl.r8 | AGGAAAAGAAATCAACCGAGA | Ma *et al.*[14] |
| mtSSU | rns.f8 | GCGCTTGACGAGTAGTTAGT | Ma *et al.*[14] |
| mtSSU | rns.r1 | AGTGGGCTACAGACGTG | This study |
| *nad4* | JM2rb | ATCTTTGGTTGCCTTTGGCA | This study |
| Non-coding region | mt.f85 | CATTATACACATTACATAATCA | Azar *et al.*[4] |
| Non-coding region | mt.f86 | CATAATATATCCTTATTCTAATGTA | This study |
| Non-coding region | mt.r63b | GTTTATAATATTGGTTATTAATTGATTTATATA | This study |
| Non-coding region | mt.r64 | GATGTATTCTATTAATTATGGTGTA | This study |
| Non-coding region | RU18.f29 | TATAGCAATTATTTATTAAGATACTTA | This study |
| Non-coding region | RU18.f30 | TACAATAGAACAATAACATATATAAG | This study |
| Non-coding region | RU18.r21 | GTAATATTAATGAATGATTTAGATTCATA | This study |
| Non-coding region | RU18.r22 | CAATATGTAAGAATTGAGGTA | This study |
| ***Nuclear genome*** | | | |
| *impdh* | 1109F | CCATCTGCAATAGTTGGTACTC | Hoffmann *et al.*[1] |
| *impdh* | 1790R | AGTAACAGGATCAGAGGGTATT | Hoffmann *et al.*[1] |
| *impdh* | IMPDH.f5 | AGGCAAATGAAATCCTCCGT | This study |
| *impdh* | IMPDH.r6 | CTGATACAGCATAAACAGCTG | This study |
| *impdh* | 2116F | TGGTGACAATACAATAGGTGATG | Hoffmann *et al.*[1] |
| *impdh* | 2859R | CATTCGTGGCGGAAGATT | Hoffmann *et al.*[1] |
| *impdh* | 1537F | CTAATCCCGCGTCAACTAAATA | Hoffmann *et al.*[1] |
| *impdh* | 2452R | CGGGTCAATGGAGGATTAAC | Hoffmann *et al.*[1] |
| *impdh* | 616F | GATAAATACCGTGGACATTCCC | Hoffmann *et al.*[1] |
| *impdh* | 2073F | CATTCAACTCCAGAGTTTCT | This study |
| *impdh* | 1461R | GCTGTATCAGAATTTGCATCTA | This study |
| *sodA* | sodA.f3 | CATTTTGTAATTGGCGTA | Azar *et al.*[4] |
| *sodA* | sodA.f4 | CTTGAACCTTATCTTTCTCA | This study |
| *sodA* | sodA.r3 | GAATCTTACATTCCATATATTTTCAA | Azar *et al.* 2022 |
| *dhps* | PS.f1 | ATCGAATGACCTTGTTCATCC | Azar *et al.* 2022 |
| *dhps* | dhps.f7 | GGTGTTCATTCATATGATTC | Ma *et al.*[14] |
| *dhps* | PS876 | ATAAATTGCAAAATTACAATCAACCAAAGT | Ma *et al.*[15] |
| ITS1-5.8S-ITS2 | ITS1.F | TTRCTGGRAAGTTGATCAAAT | Ma *et al.*[14] |
| ITS1-5.8S-ITS2 | Fun.ITS4 | TCCTCCGCTTATTGATATGC | Ma *et al.*[14] |
| ITS1-5.8S-ITS2 | Nu26S.r8 | ATTGATATGCTTAAGTTCAG | This study |
| *msg* | GK472 | TGGATCAAAAGMGAGAYTTYCCRACA | Ripamonti *et al.* [11] |
| *msg* | GK452 | AATGCACTTTCMATTGATGCT | Ripamonti *et al.*[11] |
| *msg* | GK195 | GTGTGTGTCGATGTCTGTG | Ripamonti *et al.*[11] |

Abbreviations: mtLSU, mitochondrial large subunit rRNA; mtSSU, mitochondrial small subunit rRNA*; cob*, cytochrome b; *cox2*, cytochrome c oxidase subunit 2; *cox3*, cytochrome c oxidase subunit 3; *nad4*, NADH dehydrogenase subunit 4; *impdh*, inosine monophosphate dehydrogenase; *sodA,* superoxide dismutase; *dhps*, dihydropteroate synthase; msg, major surface glycoprotein.

**Supplementary Table S2: Main conditions for PCR**

| PCR targets | Primer pairs | Amplicon size[a] | Annealing temperature for the main cycles | Extension temperature for the main cycles |
|---|---|---|---|---|
| *impdh* | 1109F-1790R[b] | 682 bp | 50 °C | 72 °C |
| | impdh.f5-impdh.r6 | 610 bp | 50 °C | 72 °C |
| | 1537F-2859R[b] | 1,323 bp | 50 °C | 72 °C |
| | 1537F-2452R | 916 bp | 50 °C | 72 °C |
| | 2116F-2859R | 744 bp | 50 °C | 72 °C |
| | 2073F-1461R[b] | 615 bp | 50 °C | 72 °C |
| | 616F-1461R | 543 bp | 50 °C | 72 °C |
| *dhps* | PS.f1-PS876[b] | 1,027 bp | 50 °C | 72 °C |
| | dhps.f7-PS876 | 830 bp | 50 °C | 72 °C |
| *sodA* | sodA.f3-sodA.r3[b] | 1,106 bp | 50 °C | 72 °C |
| | sodA.f4-sodA.r3 | 737 bp | 50 °C | 72 °C |
| ITS1-5.8S-ITS2 | ITS1.F-Fun.ITS4[b] | 633 bp | 50 °C | 72 °C |
| | ITS1.F-Nu26S.r8 | 623 bp | 50 °C | 72 °C |
| mtLSU | LSU.f1-PU3Y.f[b] | 509 bp | 50 °C | 72 °C |
| | LSU.f1-LSU.r1 | 497 bp | 50 °C | 72 °C |
| PNC | mt.f85-mt.r63b[b] | 1,028 bp | 50 °C | 72 °C |
| | mt.f86-mt.r63b | 770 bp | 50 °C | 72 °C |
| *msg* | GK472-GK452[b] | 1·7 kb | 60 °C | 72 °C |
| | GK472-GK195 | 1·4 kb | 60 °C | 72 °C |
| Mitogenome | mt.f85-rns.r1 | 4·7 kb | 45 °C | 55 °C |
| | RU18.f29-mt.r64 | 3·4 kb | 45 °C | 55 °C |
| | RU18.f30-RU18.r21 | 4·1 kb | 45 °C | 55 °C |
| | mit.f6-RU18.r22 | 4·4 kb | 45 °C | 55 °C |
| | rnl.f1 rnl.r8 | 2·3 kb | 45 °C | 65 °C |
| | cox3.f2-rnl.r2 | 5·1 kb | 45 °C | 65 °C |
| | cob.r8-JM3r | 5·4 kb | 45 °C | 65 °C |
| | JM2f-JM2rb | 4·4 kb | 45 °C | 65 °C |
| | rns.f8-JM1r | 5·2 kb | 45 °C | 65 °C |

[a]Relative to the reference sequence of the *P. jirovecii* RU7 genome.
[b]For the primary PCR while the rest is for nested or semi-nested PCR.

**Supplementary Table S3: Next-generation sequencing data sources for eight *P. jirovecii* samples**

| Patient ID | Origin (country, city) | Sampling year | Patient group | NCBI SRA accession code | References |
|---|---|---|---|---|---|
| RU7 | USA, Bethesda | Oct-1999 | Control (HIV/AIDS) | SRR1043749 | Ma *et al.*[9] |
| Pj55 | China, Chongqing | 2018 | SOT (kidney) | SRR12300271 | Chen *et al.*;[48] Cisse *et al.*[49] |
| S24 | China, Chengdu | 2015 | SOT (kidney) | CRR585179[a] | Zhao *et al.*[50] |
| Z | USA, Bethesda | Jan-1986 | Control (HIV/AIDS) | SRR5822089, SRR5825481, SRR5825487 | Ma *et al.*[9] |
| W | Denmark | Nov-1992 | Control (HIV/AIDS) | SRR1043749 | Ma *et al.*[9] |
| SE8 | Switzerland, Lausanne | Sept-2001 | Control (leukemia) | ERR135858, ERR135856, ERR135855 | Cisse *et al.*[51] |
| TJ11 | China, Tianjin | 2018-2020 | Control (leukemia) | SRR16553134, SRR16553133 | Zhao *et al.*[52] |
| TJ1 | China,Tianjin | 2018-2020 | Control (leukemia) | SRR16553134, SRR16553133 | Zhao *et al.*[52] |

[a]From the China Genome Sequence Archive at https://ngdc.cncb.ac.cn/gsa/browse/CRA008494.

**Supplementary Table S4: *P. jirovecii* isolates (n = 58) with *impdh* gene sequenced by next-generation sequencing of PCR amplicons**

| Patient IDs | Country | Minimum no. of strains[a] | IMPDH codons[b] | | | | | | | |
|---|---|---|---|---|---|---|---|---|---|---|
| | | | 225 | 253 | 261 | 285 | 289 | 308 | 323 | 343 |
| Y1 | USA | 2 | Lys | Ser | Ala | Ile | Gln | Gly | Ala | Cys |
| Y3 | USA | 2 | Lys | Ser | Ala | Ile | Gln+Arg | Gly | Ala | Cys |
| Y6 | USA | 6 | Lys | Ser | Ser | Leu | Gln | Gly | Ala | Cys |
| Y8 | USA | 2 | Lys | Ser | The | Leu | Gln+Arg | Gly | Ala | Cys |
| Y11 | USA | 5 | Lys | Ser | Ala | Leu | Gln | Gly | Ala | Cys |
| Y13 | USA | 2 | Lys | Ser | Ala | Ile | Gln | Gly | Ala | Cys |
| Y14 | USA | 2 | Lys | Ser | Ala+Thr | Ile+Leu | Gln | Gly | Ala | Cys |
| Y15 | USA | 7 | Lys | Ser | Ala | Leu | Gln | Gly | Ala | Cys |
| Y16 | USA | 7 | Lys | Ser | Ala+Thr | Ile+Leu | Gln+Arg | Gly | Ala | Cys |
| SZ5 | Switzerland | 1 | Lys | Ser | Ser | Leu | Gln | Gly | Ala | Cys |
| SZ7 | Switzerland | 1 | Lys | Ser | Ala | Leu+Ile+Val | Gln | Gly | Ala | Cys |
| SB1 | Switzerland | 2 | Lys+Asn | Ser | Ala | Leu+Ile+Val | Gln | Gly | Ala | Cys |
| SB10 | Switzerland | 2 | Lys | Ser | Thr+Ala | Leu+Ile+Val | Gln | Gly | Ala | Cys |
| SB21 | Switzerland | 3 | Lys | Ser | Thr+Ser | Leu | Gln | Gly | Ala | Cys |
| SB38 | Switzerland | 2 | Lys | Ser | The | Leu | Gln | Gly | Ala | Cys |
| G102 | Germany | 3 | Lys | Ser | Ala+Thr | Leu | Gln | Gly | Ala | Cys |
| G107 | Germany | 2 | Lys | Ser | Ala[b] | Leu | Gln | Gly | Ala | Cys |
| G130 | Germany | 3 | Lys | Ser | Ser | Leu | Gln | Gly | Ala | Cys |
| G213 | Germany | 2 | Lys | Ser | Ser | Leu | Gln | Gly | Ala | Cys |
| G349 | Germany | 2 | Lys | Ser | Ser | Leu | Gln | Gly | Ala | Cys |
| G405 | Germany | 3 | Lys+Asn | Ser | Ser+Ala | Leu+Ile+Val | Gln | Gly | Ala | Cys |
| G410 | Germany | 1 | Lys+Asn | Ser | Ala | Leu+Ile+Val | Gln | Gly | Ala | Cys |
| G665 | Germany | 1 | Lys | Ser | Ala | Leu | Gln+Arg | Gly | Ala | Cys |
| G928 | Germany | 4 | Lys | Ser | Ala | Leu | Gln | Gly | Ala | Cys |
| G979 | Germany | 1 | Lys | Ser | Ala[b] | Leu | Gln | Gly | Ala | Cys |
| J2 | Japan | 3 | Lys+Asn | Ser | Ala | Ile+Val | Gln | Gly | Ala | Cys |
| Pt3 | Japan | 1 | Lys | Ser | Ala | Leu | Gln | Gly | Ala | Cys |
| Pt4 | Japan | 1 | Lys | Ser | Ala | Leu | Leu | Gly | Ala | Cys |
| Pt6 | Japan | 2 | Lys | Ser | Ala | Leu+Val | Leu+Gln | Gly | Ala | Cys |
| Pt10 | Japan | 3 | Lys+Asn | Ser | Ala | Leu | Leu | Gly | Ala | Cys |
| D001 | Denmark | 3 | Lys+Asn | Ser | Ala | Leu+Ile+Val | Gln | Gly | Ala | Cys |
| D004 | Denmark | 2 | Lys | Ser | Ala | Val | Gln | Gly | Ala | Cys |
| D005 | Denmark | 3 | Lys | Ser | Ala | Leu | Leu+Gln | Gly | Ala | Cys |
| D014 | Denmark | 4 | Lys | Ser | Ala[b] | Leu | Gln | Gly | Ala | Cys |
| D020 | Denmark | 2 | Lys | Ser | Ala | Leu | Gln | Gly | Ala | Cys |
| D023 | Denmark | 2 | Lys | Ser | Ala | Leu | Leu | Gly | Ala | Cys |
| D024 | Denmark | 2 | Lys | Ser | Ala | Val+Leu | Gln+Leu | Gly | Ala | Cys |
| D032 | Denmark | 1 | Lys | Ser | Ala | Leu | Gln | Gly | Ala | Cys |
| D033 | Denmark | 2 | Lys | Ser | Ala | Leu | Gln | Gly | Ala | Cys |
| D042 | Denmark | 2 | Lys | Ser | Ala | Leu | Gln | Gly | Ala | Cys |
| D043 | Denmark | 3 | Lys+Asn | Ser | Ala | Val | Gln | Gly | Ala | Cys |
| D048 | Denmark | 1 | Lys | Ser | Ala | Leu | Gln | Gly | Ala | Cys |
| D049 | Denmark | 3 | Lys+Asn | Ser | Ala | Val+Leu | Gln | Gly | Ala | Cys |

| | | | | | | | | | | |
|---|---|---|---|---|---|---|---|---|---|---|
| D050 | Denmark | 3 | Lys+Asn | Ser | Ala | Leu+Ile+Val | Leu+Gln | Gly | Ala | Cys |
| GZ6 | China | 3 | Lys | Ser | Ala | Leu | Gln | Gly | Ala | Cys |
| GZ8 | China | 3 | Lys | Ser | Ala | Leu | Gln | Gly | Ala | Cys |
| GZ17 | China | 1 | Lys | Pro+Ser | Ala | Leu | Gln | Gly | Ala | Cys |
| GZ21 | China | 3 | Lys | Ser | Ala+Thr | Leu | Gln | Gly | Ala+Val | Cys+Tyr |
| GZ24 | China | 2 | Lys | Ser | Ala | Leu | Gln | Gly | Ala | Cys |
| GZ29 | China | 2 | Lys | Ser | Ala | Leu | Gln | Gly | Ala | Cys |
| GZ43 | China | 5 | Lys | Ser | Ala | Leu+Ile | Gln | Gly | Ala | Cys |
| CH4 | China | 2 | Lys | Ser | Ala | Leu | Gln | Gly | Ala | Cys |
| CH10 | China | 2 | Lys | Ser | Ala | Leu | Gln | Gly | Ala | Cys |
| CH11 | China | 1 | Lys | Ser | Ala | Leu | Gln | Gly | Ala | Cys |
| CH12 | China | 1 | Lys | Ser | Ala | Leu | Gln | Gly | Ala | Cys |
| CH17 | China | 2 | Lys | Ser | Ala | Leu | Gln | Gly | Ala | Cys |
| CH19 | China | 1 | Lys | Ser | Ala | Leu | Gln | Asp+Gly | Ala | Cys |
| CH26 | China | 2 | Lys | Ser | Ala | Leu | Gln | Gly | Ala | Cys |

[a]Based on multi-locus sequence typing with genetic markers other than *impdh* (inosine monophosphate dehydrogenase). [b]Excluding other codons with rare mutations as shown in Table 2. Mutant amino acids are highlighted in red. For sites with mixed amino acids, the dominant one is listed in the first position.

**Supplementary Table S5: Presence of multiple *impdh* alleles in *P. jirovecii* isolates from solid organ transplant patients determined by amplicon next-generation sequencing (NGS)**

| ***impdh* positions**[a] | | **No. (%) of NGS reads in isolates**[b] | | |
|---|---|---|---|---|
| Nucleotides 1020-1092-1105 | Codons 261-285-289 | G405 (Germany) | Y16 (USA) | D024 (Denmark) |
| T-C-A | Ser-Leu-Gln | 10,134 (87·1) | 0 | 0 |
| G-G-A | Ala-Val-Gln | 875 (7·5) | 21 (1·3) | 4945 (48·1) |
| T-A-A | Ser-Ile-Gln | 495 (4·3) | 0 | 0 |
| G-C-A | Ala-Leu-Gln | 65 (0·6) | 82 (5·3) | 183 (1·8) |
| G-A-A | Ala-Ile-Gln | 21 (0·2) | 829 (53·3) | 13 (0·1) |
| A-C-A | Thr-Leu-Gln | 36 (0·3) | 408 (26·2) | 11 (0·1) |
| T-G-A | Ser-Val-Gln | 13 (0·1) | 0 | 0 |
| A-A-A | Thr-Ile-Gln | 0 | 216 (13·9) | 0 |
| G-G-T | Ala-Val-Leu | 0 | 0 | 3185 (31·0) |
| G-C-T | Ala-Leu-Leu | 0 | 0 | 1927 (18·8) |
| A-G-A | Thr-Val-Gln | 0 | 0 | 7 (0·1) |
| A-C-T | Thr-Leu-Leu | 0 | 0 | 6 (0·1) |

*impdh*, inosine monophosphate dehydrogenase. [a]Non-synonymous changes (mutations) are highlighted in red. [b]All reads (150 bp) cover all three positions.

**Supplementary Table S6: Association between *P. jirovecii impdh* mutations and solid organ transplantation in Danish outbreak cases**

| Mutation[a] | Solid organ transplantation | | p value[b] |
|---|---|---|---|
| | Yes | No | |
| L285V | | | |
| Yes | 11 | 2 | |
| No | 7 | 14 | 0·004 |
| Q289L | | | |
| Yes | 8 | 1 | |
| No | 7 | 14 | 0·01 |
| L285V or Q289L | | | |
| Yes | 17 | 3 | |
| No | 7 | 14 | 0·0008 |

*impdh*, inosine monophosphate dehydrogenase.

[a]Including mutations presented as either dominant or minor variants.

[b]Chi-square test was used when appropriate, and Fisher's exact test otherwise.

When calculating p values for specific single mutations, cases with other mutations alone were excluded to ensure analytical specificity and avoid confounding.

### Supplementary Table S7: *P. jirovecii* allele assignment at three loci with low to moderate variability

***dhps*** **(dihydropteroate synthase)**

| Allele no. | Nucleotide positions | | | | | GenBank accession no. |
|---|---|---|---|---|---|---|
| | 163 | 169 | 184 | 251 | 513 | |
| 1 | A | C | G | A | A | AF139132.1 |
| 2 | A | C | G | A | G | AJ586567.1 |
| 3 | A | T | G | A | A | U66278.1 |
| 4 | G | C | G | A | A | PV684021 |
| 5 | G | T | G | A | A | U66281.1 |
| 6 | G | T | A | G | A | PV684022 |

**mtLSU (mitochondrial large subunit rRNA)**

| Allele no. | Nucleotide positions | | | | | GenBank accession no. |
|---|---|---|---|---|---|---|
| | 843 | 1006 | 1046 | 1171 | 1182-1188 | |
| 1 | A | C | A | C | CTGTAAT | PV624668 |
| 2 | A | T | A | C | CTGTAAT | PV624669 |
| 3 | AAA | C | A | C | CTGTAAT | PV624670 |
| 4 | C | C | A | C | CTGTAAT | PV624671 |
| 5 | C | C | A | C | ATTACAG | PV624672 |
| 6 | C | T | A | C | CTGTAAT | PV624673 |
| 7 | C | T | A | C | ATTACAG | PV624674 |
| 8 | T | C | A | C | CTGTAAT | PV624675 |
| 9 | T | C | A | T | CTGTAAT | PV624676 |
| 10 | T | T | A | C | CTGTAAT | PV624677 |
| 11 | C | T | G* | C | ATTACAG | PV624678 |

***sodA*** **(superoxide dismutase)**

| Allele no. | Nucleotide position | | GenBank accession no. |
|---|---|---|---|
| | 250 | 355 | |
| 1 | T | C | MW508901.1 |
| 2 | C | T | MW508902.1 |

**Supplementary Table S8: *P. jirovecii* allele assignment at two loci with high variability**

| ITSs allele no.[a] | GenBank accession no. | PNC allele no.[b] | GenBank accession no. |
|---|---|---|---|
| 1 | JQ365709 | 1 | PV614655 |
| 2 | MF980946 | 2 | PV614656 |
| 3 | JF442080 | 3 | PV614657 |
| 4 | KC470776 | 4 | PV614658 |
| 5 | KC470798 | 5 | PV614659 |
| 6 | KC470788 | 6 | PV614660 |
| 7 | MF980974 | 7 | PV614661 |
| 8 | U07220 | 8 | PV614662 |
| 9 | JQ365723 | 9 | PV614663 |
| 10 | JQ365725 | 10 | PV614664 |
| 11 | JQ365748 | 11 | PV614665 |
| 12 | AB481406 | 12 | PV614666 |
| 13 | AB481411 | 13 | PV614667 |
| 14 | JQ365710 | 14 | PV614668 |
| 15 | JQ365728 | 15 | PV614669 |
| 16 | AB469817 | 16 | PV614670 |
| 17 | AB481410 | 17 | PV614671 |
| 18 | AB481413 | 18 | PV614672 |
| 19 | JQ365708 | 19 | PV614673 |
| 20 | JQ365715 | 20 | PV614674 |
| 21 | JQ365718 | 21 | PV614675 |
| 22 | KC470795 | 22 | PV614676 |
| 23 | MF989107 | 23 | PV614677 |
| 24 | AB481412 | 24 | PV614678 |
| 25 | JF442094 | 25 | PV614679 |
| 26 | JF442099 | 26 | PV614680 |
| 27 | JQ365707 | 27 | PV614681 |
| 28 | JQ365712 | 28 | PV614682 |
| 29 | JQ365717 | 29 | PV614683 |
| 30 | JQ365724 | 30 | PV614684 |
| 31 | JQ365730 | 31 | PV614685 |
| 32 | JQ365731 | 32 | PV614686 |
| 33 | JQ365737 | 33 | PV614687 |
| 34 | JQ365740 | 34 | PV614688 |
| 35 | JQ365741 | 35 | PV614689 |
| 36 | JQ365746 | 36 | PV614690 |
| 37 | KC470771 | 37 | PV614691 |
| 38 | KC470772 | 38 | PV614692 |
| 39 | KC470773 | 39 | PV614693 |
| 40 | KC470774 | 40 | PV614694 |
| 41 | KC470775 | 41 | PV614695 |
| 42 | KC470785 | 42 | PV614696 |
| 43 | KC470787 | 43 | PV614697 |
| 44 | MF989229 | 44 | PV614698 |
| 45 | MF989230 | 45 | PV614699 |
| 46 | MF989231 | 46 | PV614700 |
| 47 | MF989232 | 47 | PV614701 |
| 48 | MF989233 | 48 | PV614702 |
| 49 | MF989234 | 49 | PV614703 |
| 50 | U07221 | 50 | PV614704 |
| 51 | U07226 | 51 | PV614705 |
| 52 | MK300654 | 52 | PV614706 |
| 53 | MK300659 | 53 | PV614707 |
| 54 | MK300656 | 54 | PV614708 |
| 55 | MK300657 | 55 | PV614709 |
| 56 | MK300658 | 56 | PV614710 |
| 57 | MK300655 | 57 | PV614711 |
| 58 | MK300660 | 58 | PV614712 |

| | | | |
|---|---|---|---|
| 59 | MK300661 | 59 | PV614713 |
| 60 | MK300662 | 60 | PV614714 |
| 61 | MK300663 | 61 | PV614715 |
| 62 | MK300664 | 62 | PV614716 |
| 63 | MW646092 | 63 | PV614717 |
| 64 | PV614139 | 64 | PV614718 |
| 65 | PV614140 | 65 | PV614719 |
| 66 | PV614141 | 66 | PV614720 |
| 67 | PV614142 | 67 | PV614721 |
| 68 | PV614143 | | |
| 69 | PV614144 | | |
| 70 | PV614145 | | |

[a]ITSs refers to the region spanning the nuclear internal transcribed spacer 1 (ITS1), 5.8S rRNA and internal transcribed spacer 2 (ITS2) of the rRNA operon, also known as ITS1-5.8S-ITS2.[24] Alleles 1 to 62 are reported by Xue et al. 2019; [24] allele 63 is reported by Azar et al. 2022.[4] [b]PNC refers to a polymorphic non-coding region of the mitochondrial genome. Alleles 1 to 22 are identical, within the overlapping region, to the short versions of PNC **t**ypes 1 to 22 described by Azar et al. 2022.[4]

**Supplementary Table S9: Dominant genotype profiles of 43 *P. jirovecii* isolates based on five genetic loci***

| Isolate ID | Origin (country, city) | *dhps* allele | sodA allele | mtLSU allele | PNC allele | ITSs allele |
|---|---|---|---|---|---|---|
| CH26 | China, Guangzhou | 1 | 1 | 1 | 45 | 64 |
| GR11 | Germany, Zurich | 1 | 1 | 4 | 5 | 1 |
| Z | USA, Bethesda | 1 | 1 | 4 | 5 | 53 |
| D013 | Denmark, Copenhagen | 1 | 1 | 4 | 6 | 1 |
| CH51 | China, Guangzhou | 1 | 1 | 6 | 61 | 22 |
| CH04 | China, Guangzhou | 1 | 1 | 7 | 51 | 1 |
| A | USA, Bethesda | 1 | 1 | 8 | 17 | 10 |
| D014 | Denmark, Copenhagen | 1 | 1 | 8 | 24 | 41 |
| D001 | Denmark, Copenhagen | 1 | 1 | 8 | 44 | 17 |
| GZ43 | China, Guangzhou | 1 | 1 | 9 | 51 | 1 |
| GZ32 | China, Guangzhou | 1 | 1 | 10 | 68 | 71 |
| D020 | Denmark, Copenhagen | 1 | 2 | 1 | 18 | 40 |
| D040 | Denmark, Copenhagen | 1 | 2 | 1 | 38 | 22 |
| GZ02 | China, Guangzhou | 1 | 2 | 1 | 41 | 1 |
| D012 | Denmark, Copenhagen | 1 | 2 | 1 | 50 | 39 |
| D044 | Denmark, Copenhagen | 1 | 2 | 4 | 33 | 37 |
| CH30 | China, Guangzhou | 1 | 2 | 4 | 66 | 1 |
| CH10 | China, Guangzhou | 1 | 2 | 5 | 12 | 53 |
| D048 | Denmark, Copenhagen | 1 | 2 | 8 | 23 | 43 |
| GZ37 | China, Guangzhou | 1 | 2 | 8 | 57 | 1 |
| SZ7 | Switzerland, Zurich | 2 | 1 | 1 | 30 | 5 |
| GZ14 | China, Guangzhou | 2 | 1 | 1 | 53 | 62 |
| GZ24 | China, Guangzhou | 2 | 1 | 1 | 55 | 1 |
| GZ03 | China, Guangzhou | 2 | 1 | 2 | 40 | 16 |
| GZ42 | China, Guangzhou | 2 | 1 | 6 | 49 | 9 |
| CH12 | China, Guangzhou | 2 | 1 | 6 | 52 | 4 |
| GZ29 | China, Guangzhou | 2 | 1 | 6 | 56 | 70 |
| CH19 | China, Guangzhou | 2 | 1 | 7 | 13 | 53 |
| CH16 | China, Guangzhou | 2 | 1 | 7 | 54 | 59 |
| D042 | Denmark, Copenhagen | 2 | 1 | 8 | 4 | 22 |
| GZ08 | China, Guangzhou | 2 | 1 | 9 | 7 | 69 |
| D039 | Denmark, Copenhagen | 2 | 2 | 1 | 34 | 12 |
| CH11 | China, Guangzhou | 2 | 2 | 1 | 59 | 67 |
| D032 | Denmark, Copenhagen | 2 | 2 | 3 | 23 | 17 |
| CH17 | China, Guangzhou | 2 | 2 | 4 | 60 | 10 |
| GZ01 | China, Guangzhou | 2 | 2 | 4 | 63 | 32 |
| GZ16 | China, Guangzhou | 2 | 2 | 6 | 64 | 53 |
| D033 | Denmark, Copenhagen | 2 | 2 | 8 | 18 | 42 |
| SE8 | Switzerland, Lausanne | 3 | 1 | 8 | 34 | 4 |
| R68 | USA, Bethesda | 4 | 2 | 4 | 46 | 5 |
| W | Denmark, Copenhagen | 4 | 2 | 6 | 28 | 1 |
| RU7 | USA, Bethesda | 5 | 1 | 1 | 2 | 17 |
| S567 | USA, San Francisco | 5 | 1 | 1 | 31 | 6 |

*Including dominant alleles alone and excluding isolates with any sites not successfully sequenced. The sequence and allele assignment for each isolate are provided in the Supplementary Tables S7-S8. *dhps*, dihydropteroate synthase; *sodA*, superoxide dismutase; mtLSU, mitochondrial large subunit rRNA; ITSs, nuclear internal transcribed spacer 1 (ITS1), 5.8S rRNA and ITS2 of the rRNA operon (ITS1-5.8S-ITS2); PNC, polymorphic non-coding region of the mitogenome.

**Supplementary Table S10: Discriminatory index for multi-locus sequence typing schemes based on various combinations of 5 genetic markers[a]**

| Genetic markers | No. of genotypes[a] | Discrimination index[b] |
|---|---|---|
| *sodA* alone | 2 | 0·49883 |
| *dhps* alone | 5 | 0·6179 |
| mtLSU alone | 10 | 0·8439 |
| ITSs alone | 28 | 0·9502 |
| PNC alone | 39 | 0·9956 |
| *dhps* + *sodA* + mtLSU + ITSs | 42 | 0·9989 |
| PNC + ITSs | 42 | 0·9989 |
| *dhps* + *sodA* + mtLSU + PNC | 43 | 1·0000 |
| All 5 loci | 43 | 1·0000 |

*dhps*, dihydropteroate synthase (with mutations at codons 55 and/or 57 in 5 isolates as detailed in Supplementary Table S7); *sodA,* superoxide dismutase; mtLSU, mitochondrial large subunit rRNA; ITSs, nuclear internal transcribed spacer 1 (ITS1), 5.8S rRNA and ITS2 of the rRNA operon (ITS1-5.8S-ITS2); PNC, polymorphic non-coding region of the mitogenome. [a]See the detail in Supplementary Tables S7-S8. [b]Calculated using data in Supplementary Table S9 according to the method by Hunter et al.[26]

**Supplementary Table S11: Coinfection of two distinct *P. jirovecii* strains with different *impdh* mutations (C1092G/L285V and A1105T/Q289L) in a transplant patient**

| Genes | Nucleotide Positions | D024 strains[a] *a* | *b* | Ratio *a*/*b* | D043 | D007 | Pt6 |
|---|---|---|---|---|---|---|---|
| *impdh* | 1020 | G | G | 1/1 | G | G | G |
| | 1092 | G | C | 1·6/1 | G | C | C |
| | 1105 | A | T | 1·2/1 | A | T | T |
| | 1329 | T | A | 2·2·1 | T | A | A |
| | 1386 | C | A | 2·2/1 | C | A | A |
| *dhps* allele | | 1 | 2 | 1·4/1 | 1 | 2 | 2 |
| *soda* allele | | 1 | 2 | 1·7/1 | 1 | 2 | 1 |
| mtLSU allele | | 6 | 8 | 1·3/1 | 6 | 8 | 8 |
| PNC allele | | 20 | 3 | 1·4/1 | 20 | 3 | 3 |
| ITSs allele | | 4 | 5 | 1·3/1 | 4 | 5 | 17 |
| mtDNA[b] | | PV684663 | PV684664 | 1·1/1 | PV684663 | PV684664 | PV684670 |

[a]The ratio of the two alleles at each position was calculated with the number of Illumina reads. The dominant strain *a* in D024 (from Denmark) shared an identical MLST profile with strain D043 (Denmark) while the minor strain *b* shared an identical profile with strain D007 (Denmark) but not strain Pt6 (Japan) despite the presence of the same mutation (A1105T/Q289L) between Pt6 and D007/D024(*b*). Non-synonymous changes are highlighted in red. Identical genotypes are indicated in the same color. Details for MLST allele profiles are available in Supplementary Tables S7-S8. Similar coinfection occurred in patient Y16 from USA, which included a minor strain sharing an identical MLST profile and an identical mutation (G1020A/A261T) with the dominant strain in another patient (Y14) from the same outbreak as described by Azar et al.[4]
[a]identical sequences were assigned the same accession nos. PV684663 or PV684664.
*impdh*, inosine monophosphate dehydrogenase.

**Supplementary Table S12. Correlation of mitogenome (mtDNA) data with *impdh* mutations and multi-locus sequence typing (MLST) profiles in 40 selected *P. jirovecii* isolates**

| Patient ID | Country | Patient group | *impdh* mutations | MLST profile[a] | mtDNA GenBank accession nos. | Notes for mtDNA |
|---|---|---|---|---|---|---|
| SZ7 | Switzerland | Control | No | 2-1-1-30-5 | MH010446 | Unique |
| SE8 | Switzerland | Control | No | 3-1-8-34-4 | PV684658 | Unique |
| G410 | Germany | Control | No | 1-1-4-5-1 | MH010440 | Unique |
| D001 | Denmark | Control | No | 1-1-8-44-17 | PV684660 | Unique |
| W | Denmark | Control | No | 4-2-6-28-1 | JX855936 | Unique |
| TJ11 | China | Control | No | n-1-1-37-n | PV684659 | Unique |
| CH26 | China | Control | No | 1-1-1-45-64 | MH010438 | Unique |
| 1368 | USA | Control | No | n-n-6-14-n | JX855938 | Unique |
| S567 | USA | Control | No | 5-1-1-31-6 | PV684657 | Unique |
| RU7 | USA | Control | No | 5-1-1-2-17 | NC_020331 | Unique |
| Z | USA | Control | No | 1-1-4-5-53 | PV684655 | Unique |
| A | USA | Control | No | 1-1-8-17-10 | PV684656 | Unique |
| R68 | USA | Control | No | 4-2-4-46-5 | JX855937 | Unique |
| Y6 | USA | SOT (kidney) | A261S | 1-2-8-3-53 | MW554494[b] | Unique |
| Pt2 | Japan | SOT (kidney) | L285V | n-1-9-1-53 | PV684669 | Unique |
| Pt6 | Japan | SOT (kidney) | Q289L | 2-1-9-3-17 | PV684670 | Identical |
| Pt10 | Japan | SOT (kidney) | Q289L | 2-1-9-3-17 | PV684670 | |
| Y13 | USA | SOT (kidney) | L285I | 2-1-9-3-22 | MW652730[b] | All 4 are identical in overlapping regions (only partial mtDNA of 11-21 kb in Y3 and Y13) |
| Y1 | USA | SOT (kidney) | L285I | 2-1-9-3-22 | MW530525[b] | |
| Y3 | USA | SOT (kidney) | L285I | 2-1-9-3-22 | MW652729 | |
| Y16 | USA | SOT (kidney) | L285I | 2-1-9-3-22 | MW530525 | |
| D022 | Denmark | SOT (liver) | No | 1-2-6-25-6 | PV684667 | All 4 are identical except for a single base deletion for a C homopolymer in D041 |
| D041 | Denmark | SOT (kidney) | No | 1-2-6-25-6 | PV684668 | |
| D008 | Denmark | SOT (kidney) | No | 1-2-6-25-6 | PV684667 | |
| D027 | Denmark | SOT (liver) | No | 1-2-6-25-6 | PV684667 | |
| D007 | Denmark | SOT (kidney) | Q289L | 2-2-9-3-5 | PV684664 | Identical |
| D050 | Denmark | SOT (liver) | Q289L | 2-2-9-3-5 | PV684664 | |
| D024b[c] | Denmark | SOT (kidney) | Q289L | 2-2-9-3-5 | PV684664 | |
| D024a[c] | Denmark | SOT (kidney) | L285V | 1-1-6-20-4 | PV684663 | Identical except for a deletion of one AGT repeat unit and an insert of a single base for an A homopolymer in D049 |
| D043 | Denmark | SOT (kidney) | L285V | 1-1-6-20-4 | PV684661 | |
| D049 | Denmark | SOT (liver) | L285V | 1-1-6-20-4 | PV684662 | |
| Pj55 | China | SOT (kidney) | L285I | 2-2-9-3-6 | PV684665 | Identical except for variation in the copy number of tandem repeats at 6 positions and SNPs in 3 positions (32871, 33319 and 33371 in Pj55). These 3 positions in S24 contain a mixture of two alleles, with the minor one identical to that of Pj55. |
| S24 | China | SOT (kidney) | A261T | 2-2-9-3-n | PV684666 | |
| SB24 | Switzerland | SOT (kidney) | A261T | 1-2-7-1-1 | PV684671 | Identical except for a single base deletion for a homopolymer in SW12 and SW24 |
| SB12 | Switzerland | SOT (kidney) | A261T | 1-2-7-1-1 | PV684671 | |
| Y14 | USA | SOT (kidney) | A261T | 1-2-7-1-1 | MW530526 | |
| Y8 | USA | SOT (kidney) | A261T | 1-2-7-1-1 | MW530526 | |
| G405 | Germany | SOT (kidney) | A261S | 1-2-1-18-53 | MH010441 | |

| | | | | | | |
|---|---|---|---|---|---|---|
| SZ1 | Switzerland | SOT (kidney) | A261S | 1-2-1-18-53 | MH010444 | Identical except for a single base deletion for a T homopolymer in G405 |
| SZ4 | Switzerland | SOT (kidney) | A261S | 1-2-1-18-53 | MH010445 =MH010444 | |

[a]Allele assignment is available in Supplementary Tables S7-S8. Undetermined genotypes are indicated by "n". Samples with identical MLST profiles and mtDNA sequences (excluding variations in tandem repeat copy numbers) are indicated by the same colors. [b] Only these three were partial (11-23 kb), whereas all others were complete (33-35 kb). [c]Two strains ientified from the same patient D024 by MLST and mtDNA sequence (see more details in Supplementary Table S11). *impdh*, inosine monophosphate dehydrogenase.

**Supplementary Table S13: Correlation between *impdh* mutations, multi-locus sequence typing (MLST) profiles and restriction fragment length polyphism (RFLP) patterns of dominant *P. jirovecii* strains**

| Patient ID[a] | Country and city | Patient group | *impdh* variants | MLST profile[b] | RFLP pattern | Reference |
|---|---|---|---|---|---|---|
| SB2 | Switzerland, Bern | Control | Wild type | 1-1-1-58-1 | Unique | This study |
| SB3 | Switzerland, Bern | Control | Wild type | 1-1-8-37-1 | Unique | This study |
| SB6 | Switzerland, Bern | SOT (kidney) | Wild type | 1-1-8-34-6 | Unique | This study |
| SB5 | Switzerland, Bern | SOT (kidney) | H108Y | 2-1-1-32-17 | Identical | This study |
| SB7 | Switzerland, Bern | SOT (kidney) | H108Y | 2-1-1-32-17 | | This study |
| SB8 | Switzerland, Bern | SOT (kidney) | H108Y | 2-1-1-32-17 | | This study |
| SB9 | Switzerland, Bern | SOT (kidney) | H108Y | 2-1-1-32-17 | | This study |
| SB10 | Switzerland, Bern | SOT (kidney) | A261T | 1-2-7-1-1 | Identical | This study |
| SB11 | Switzerland, Bern | SOT (kidney) | A261T | 1-2-7-1-1 | | This study |
| SB12 | Switzerland, Bern | SOT (kidney) | A261T | 1-2-7-1-1 | | This study |
| SB16 | Switzerland, Bern | SOT (kidney) | A261T | 1-2-7-1-1 | | This study |
| SB20 | Switzerland, Bern | SOT (kidney) | A261T | 1-2-7-1-1 | | This study |
| SB24 | Switzerland, Bern | SOT (kidney) | A261T | 1-2-7-1-1 | | This study |
| SB37 | Switzerland, Bern | SOT (kidney) | A261T | 1-2-7-1-1 | | This study |
| SB39 | Switzerland, Bern | SOT (kidney) | A261T | 1-2-7-1-1 | | This study |
| SB40 | Switzerland, Bern | SOT (kidney) | A261T | 1-2-7-1-1 | | This study |
| SZ7 | Switzerland, Zurich | Control | Wild type | 2-1-1-30-5 | Unique | Sassi et al.[13] |
| SZ1 | Switzerland, Zurich | SOT (kidney) | A261S | 1-2-1-18-53 | Identical | Sassi et al.[13] |
| SZ2 | Switzerland, Zurich | SOT (kidney) | A261S | 1-2-1-18-53 | | Sassi et al.[13] |
| SZ3 | Switzerland, Zurich | SOT (kidney) | A261S | 1-2-1-18-53 | | Sassi et al.[13] |
| SZ4 | Switzerland, Zurich | SOT (kidney) | A261S | 1-2-1-18-53 | | Sassi et al.[13] |
| SZ5 | Switzerland, Zurich | SOT (kidney) | A261S | 1-2-1-18-53 | | Sassi et al.[13] |
| G752 | Germany, Munich | SOT (kidney) | A261S | 1-2-1-18-53 | | Ripamonti et al.[11] |
| G380 | Germany, Munich | SOT (kidney) | A261S | 1-2-1-18-53 | | Ripamonti et al.[11] |
| G682 | Germany, Munich | SOT (kidney) | A261S | 1-2-1-18-53 | | Ripamonti et al.[11] |
| G908 | Germany, Munich | SOT (kidney) | A261S | 1-2-1-18-53 | | Ripamonti et al.[11] |
| G371 | Germany, Munich | SOT (kidney) | A261S | 1-2-1-18-53 | | Ripamonti et al.[11] |
| G534 | Germany, Munich | SOT (kidney) | A261S | 1-2-1-18-53 | | Ripamonti et al.[11] |
| G549 | Germany, Munich | SOT (kidney) | A261S | 1-2-1-18-53 | | Ripamonti et al.[11] |
| G643 | Germany, Munich | SOT (kidney) | A261S | 1-2-1-18-53 | | Ripamonti et al.[11] |
| G349 | Germany, Munich | SOT (kidney) | A261S | 1-2-1-18-53 | | Ripamonti et al.[11] |
| G213 | Germany, Munich | SOT (kidney) | A261S | 1-2-1-18-53 | | Ripamonti et al.[11] |
| G130 | Germany, Munich | SOT (kidney) | A261S | 1-2-1-18-53 | | Ripamonti et al.[11] |
| G405 | Germany, Munich | SOT (kidney) | A261S | 1-2-1-18-53 | | Ripamonti et al.[11] |
| G102 | Germany, Munich | SOT (kidney) | A261S[c] | n-2-1-n-n | | Ripamonti et al.[11] |
| G665 | Germany, Munich | Control | Wild type | n-n-8-20-n | Unique | Ripamonti et al.[11] |
| G410 | Germany, Munich | Control | Wild type | 1-1-4-5-1 | Unique | Ripamonti et al.[11] |
| G181 | Germany, Munich | Control | Wild type | n-2-4-10-n | Unique | Ripamonti et al.[11] |
| G107 | Germany, Munich | Control | Wild type | n-2-1-6-n | Unique | Ripamonti et al.[11] |
| G928 | Germany, Munich | Control | Wild type | n-2-4-8-n | Unique | Ripamonti et al.[11] |
| G979 | Germany, Munich | Control | Wild type | n-n-1-38-n | Unique | Ripamonti et al.[11] |
| J1 | Japan, Nagoya | SOT (kidney) | L285I | 2-2-9-3-22 | Identical | Ripamonti et al.[11] |

| | | | | | | |
|---|---|---|---|---|---|---|
| J2 | Japan, Nagoya | SOT (kidney) | L285I | 2-2-9-3-22 | | Ripamonti et al.[11] |
| J5 | Japan, Nagoya | SOT (kidney) | L285I | 2-2-9-3-22 | | Ripamonti et al.[11] |
| J8 | Japan, Nagoya | SOT (kidney) | L285I | 2-2-n-3-22 | Unique[d] | Ripamonti et al.[11] |
| D002 | Denmark, Copenhagen | SOT (liver) | L285I | 1-2-6-25-1 | Identical | Rostved et al.[12] |
| D053 | Denmark, Copenhagen | Control | L285I | 1-2-6-25-1 | | Rostved et al.[12] |
| D008 | Denmark, Copenhagen | SOT (kidney) | L285I | 1-2-6-25-6 | Identical | Rostved et al.[12] |
| D010 | Denmark, Copenhagen | SOT (kidney) | L285I | 1-2-6-25-6 | | Rostved et al.[12] |
| D022 | Denmark, Copenhagen | SOT (kidney) | L285I | 1-2-6-25-6 | | Rostved et al.[12] |
| D027 | Denmark, Copenhagen | SOT (liver) | L285I | 1-2-6-25-6 | | Rostved et al.[12] |
| D041 | Denmark, Copenhagen | SOT (kidney) | L285I | 1-2-6-25-6 | | Rostved et al.[12] |
| D003 | Denmark, Copenhagen | SOT (kidney) | L285V | 1-1-6-20-4 | Identical | Rostved et al.[12] |
| D028 | Denmark, Copenhagen | SOT (kidney) | L285V | 1-1-6-20-4 | | Rostved et al.[12] |
| D043 | Denmark, Copenhagen | SOT (kidney) | L285V | 1-1-6-20-4 | | Rostved et al.[12] |
| D047 | Denmark, Copenhagen | SOT (kidney) | L285V | 1-1-6-20-4 | | Rostved et al.[12] |
| D049 | Denmark, Copenhagen | SOT (liver) | L285V | 1-1-6-20-4 | | Rostved et al.[12] |
| D051 | Denmark, Copenhagen | SOT (liver) | L285V | 1-1-6-20-4 | | Rostved et al.[12] |
| D005 | Denmark, Copenhagen | SOT (liver) | Q289L | 2-2-9-3-5 | Identical | Rostved et al.[12] |
| D007 | Denmark, Copenhagen | SOT (kidney) | Q289L | 2-2-9-3-5 | | Rostved et al.[12] |
| D009 | Denmark, Copenhagen | SOT (kidney) | Q289L | 2-2-9-3-5 | | Rostved et al.[12] |
| D016 | Denmark, Copenhagen | SOT (liver) | Q289L | 2-2-9-3-5 | | Rostved et al.[12] |
| D017 | Denmark, Copenhagen | SOT (kidney) | Q289L | 2-2-9-3-5 | | Rostved et al.[12] |
| D050 | Denmark, Copenhagen | SOT (liver) | Q289L | 2-2-9-3-5 | | Rostved et al.[12] |
| D001 | Denmark, Copenhagen | Control | Wild type | 1-1-8-44-17 | Unique | Rostved et al.[12] |
| D032 | Denmark, Copenhagen | Control | Wild type | 2-2-3-23-17 | Unique | Rostved et al.[12] |
| D033 | Denmark, Copenhagen | Control | Wild type | 2-2-8-18-42 | Unique | Rostved et al.[12] |
| D040 | Denmark, Copenhagen | Control | Wild type | 1-2-1-38-22 | Unique | Rostved et al.[12] |
| D042 | Denmark, Copenhagen | Control | Wild type | 2-1-8-4-22 | Unique | Rostved et al.[12] |
| D044 | Denmark, Copenhagen | Control | Wild type | 1-2-4-33-37 | Unique | Rostved et al.[12] |
| Y1 | USA, New Haven | SOT (kidney) | L285I | 2-1-9-3-22 | Identical | Azar et al. [4] |
| Y3 | USA, New Haven | SOT (kidney) | L285I | 2-1-9-3-22 | | Azar et al. [4] |
| Y13 | USA, New Haven | SOT (kidney) | L285I | 2-1-9-3-22 | | Azar et al. [4] |
| Y16 | USA, New Haven | SOT (kidney) | L285I | 2-1-9-3-22 | Unique[d] | Azar et al. [4] |
| Y14 | USA, New Haven | SOT (kidney) | L285I | 1-2-7-1-1 | Identical | Azar et al. [4] |
| Y8 | USA, New Haven | SOT (kidney) | A261T | 1-2-7-1-1 | | Azar et al. [4] |
| Y6 | USA, New Haven | SOT (kidney) | A261S | 1-2-8-3-53 | Unique | Azar et al. [4] |
| Y11 | USA, New Haven | SOT (kidney) | Wild type | 1-1-6-25-63 | Unique | Azar et al. [4] |
| Y15 | USA, New Haven | SOT (kidney) | Wild type | 1-2-1-37-1 | Unique | Azar et al. [4] |

[a]Including only samples analyzed by both RFLP and *P. jirovecii impdh* sequencing. Samples with identical MLST profiles and RFLP patterns are indicated by the same colors. [b]Undetermined genotypes are indicated by "n". [c]Containing a mixture of A261S and wild type. [d]Only these two samples showed a distinct RFLP pattern while sharing an identical MLST profile with the others. *impdh*, inosine monophosphate dehydrogenase

**Supplementary Table S14: Genotype profiles of *P. jirovecii* isolates with no *impdh* mutations in 12 organ transplant patients.**

| No. of isolates | Countries (sample ID) | *impdh* variant types[a] | MLST profiles at 5 genetic loci[b] | No. of isolates with mitogenome available |
|---|---|---|---|---|
| 1 | Denmark (D031) | 1 | 2-2-2-43-1 | 0 |
| 5 | Denmark (D008, D010 D022, D027, D041) | 2 | 1-2-6-25-6 | 4[c] |
| 1[d] | Denmark (D002) | 2 | 1-2-6-25-1 | 0 |
| 1 | USA (Y11) | 2 | 1-1-6-14-63 | 1 |
| 1 | USA (Y15) | 8 | 1-2-1-18-1 | 1 |
| 1 | Switzerland (SB6) | 7 | 1-1-8-34-6 | 0 |
| 1 | Switzerland (SB35) | 9 | n-2-8-26-1 | 0 |
| 1 | Switzerland (SB13) | 18 | n-2-1-18-n | 0 |

*impdh*, inosine monophosphate dehydrogenase; MLST, multi-locus sequence typing.
[a]No missense mutations (either as dominant or minor populations) throughout the full-length gene (except for unsequenced 900-bp 5' end in SB13).
[b]See allele assignment in Supplementary Tables S7-S8. n, undetermined genotype.
[c]Shared an identical full mitogenome sequence (D010 not sequenced).
[d]This isolate shared an identical MLST profile with the isolate from a non-transplant Danish patient (D053).

**Supplementary Table S15: *P. jirovecii* samples and their common mutations in the *impdh* gene.**

| Country | Transplant group (n = 96) | | | | | | | Control group (n = 67) | | | | | | |
|---|---|---|---|---|---|---|---|---|---|---|---|---|---|---|
| | H108Y | A261T | A261S | L285I | L285V | Q289L | *Total* | H108Y | A261T | A261S | L285I | L285V | Q289L | *Total* |
| USA | 0 | 2 | 1 | 5 | 0 | 0 | 7*/9 | 0 | 0 | 0 | 0 | 0 | 0 | 0/6 |
| Denmark | 0 | 0 | 0 | 0 | 10 | 7 | 17/24 | 0 | 0 | 0 | 1 | 2 | 1 | 3/18 |
| Germany | 0 | 0 | 13 | 1 | 1 | 0 | 13*/13 | 0 | 0 | 0 | 1 | 1 | 1 | 2*/6 |
| Switzerland | 4 | 16 | 6 | 1 | 1 | 0 | 25*/28 | 0 | 0 | 0 | 1 | 1 | 0 | 2*/6 |
| Japan | 0 | 0 | 2 | 8 | 2 | 9 | 19*/20 | 0 | 0 | 0 | 0 | 0 | 0 | 0/1 |
| China | 0 | 1 | 0 | 1 | 0 | 0 | 2/2 | 0 | 0 | 1 | 1 | 0 | 0 | 2/30 |
| *Total* | *4* | *19* | *22* | *16* | *14* | *16* | *83/96* | *0* | *0* | *1* | *4* | *4* | *2* | *9/67* |

*impdh*, inosine monophosphate dehydrogenase. *Including one or two patients infected with *P. jirovecii* isolates containing two types of mutations at the same sites or mutations at multiple sites either as dominant or minor populations.

**Supplementary Table S16: Frequency of common i*mpdh* mutations in *P. jirovecii* samples from transplant patients.**

| **Country** | **A261S** | **A261T** | **H108Y** | **L285I** | **L285V** | **Q289L** | **Other** | **Total** |
|---|---|---|---|---|---|---|---|---|
| China | 0 (0·0) | 1 (50·0) | 0 (0·0) | 1 (50·0) | 0 (0·0) | 0 (0·0) | 0 (0·0) | *2* |
| Denmark | 0 (0·0) | 0 (0·0) | 0 (0·0) | 0 (0·0) | 10 (41·7) | 7 (29·2) | 7 (29·2) | *24* |
| Germany | 13 (86·7) | 0 (0·0) | 0 (0·0) | 1 (6·7) | 1 (6·7) | 0 (0·0) | 0 (0·0) | *15** |
| Japan | 2 (9·1) | 0 (0·0) | 0 (0·0) | 8 (36·4) | 2 (9·1) | 9 (40·9) | 1 (4·6) | *22** |
| Switzerland | 6 (19·4) | 16 (51·6) | 4 (12·9) | 1 (3·2) | 1 (3·2) | 0 (0·0) | 3 (9·7) | *31** |
| USA | 1 (10·0) | 2 (20·0) | 0 (0·0) | 5 (50·0) | 0 (0·0) | 0 (0·0) | 2 (20·0) | *10** |
| *Total* | *22* | *19* | *4* | *16* | *14* | *16* | *13* | *104* |

Values in the cells are the numbes (%) of patients carrying *P. jirovecii impdh* mutations as shown or no mutations (denoted as "Other, which includes wild-type and other synonymous and nonsynonymous changes) based on the data from transplant patients included in Table S15 above. *Patients carrying more than one mutation were counted for each mutation, causing the total number of samples (n = 104) to be larger than the total number of transplant patients (n = 96) as shown above in Table S15. The distributions of these mutations differ significantly across ountries (Likelihood Ratio test: $p < 0.0001$). *impdh*, inosine monophosphate dehydrogenase.

**Supplementary Table S17: *P. jirovecii impdh* mutation shifts in transplant patients across different outbreaks**

| | IMPDH mutations | No. of strain (strain designation*) |
|---|---|---|
| Japan (one hospital, three outbreaks) | | |
| 1st outbreak 2005 (n = 8) | L285I[a] | 1 (JP18) |
| 2nd outbreak 2011 (n = 2) | A261S | 1 (JP35) |
| 2nd outbreak 2011 (n = 1) | Wild type (*impdh*-4) | 1 |
| 3rd outbreaks 2013-2015 (n = 9) | Q289L[a] | 1 (JP38) |
| Switzerland (two hospitals, three outbreaks) | | |
| 1st Bern 2005 (n = 4) | H108Y | 1 (SW9) |
| 1st Bern 2005 (n = 1) | Wild type (*impdh*-17) | 1 |
| 2nd Bern 2009-2012 (n = 16) | A261T[b] | 1 (EU8) |
| 2nd Bern 2009-2012 (n = 2) | Wild type (*impdh*-9/18) | 2 |
| 3rd Zurich 2007 (n = 5) | A261S | 1 (GS5) |
| Denmark (one hospital, one outbreak) | | |
| Sept 2007-May 2009 (n = 5) | Wild type (*impdh*-2[c]) | 1 |
| June 2009-July 2009 (n = 2[d]) | Wild type (*impdh*-2[c]) | 1 |
| June 2009-Nov 2010 (n = 11[d]) | L285V[e] | 1 (DK2) |
| Oct 2009-July 2010 (n = 8[d]) | Q289L[d] | 1 (DK3) |
| Nov 2009 (n = 1) | Wild type (*impdh*-1) | 1 |

*See detail for mutant strain designation in Table 4 and Figure 3. [a]Including one case containing L285V as an additional minor population (in J2 and Pt6). [b]Including two cases containing additional mutations A261S (in SB21) or L285I and L285V (in SB10) as minor populations. [c]Different strains based on MLST profiles. [d]Including one non-transplant control. [e]Including one case (D024) containing Q289L as an additional minor population. [f]Including one case (D050) containing L285I and L285V as additional minor populations. *impdh*, inosine monophosphate dehydrogenase.

**Supplementary Table S18: Predicted mutation effects on IMPDH protein stability (versus wild-type) based on multiple methods.**

| Model | DDMut | DynaMut2 | DDGun | Eris | DUET | MUpro | mCSM | iStable | Consensus |
|---|---|---|---|---|---|---|---|---|---|
| H108Y | Stabilizing | Stabilizing | Stabilizing | Destabilizing | Stabilizing | Destabilizing | Stabilizing | Stabilizing | Stabilizing |
| A261T | Destabilizing | Destabilizing | Destabilizing | Destabilizing | Destabilizing | Destabilizing | Destabilizing | Destabilizing | Destabilizing |
| A261S | Destabilizing | Destabilizing | Destabilizing | Destabilizing | Destabilizing | Destabilizing | Destabilizing | Destabilizing | Destabilizing |
| L285I | Destabilizing | Destabilizing | Stabilizing | Stabilizing | Destabilizing | Destabilizing | Destabilizing | Destabilizing | Destabilizing |
| L285V | Destabilizing | Destabilizing | Destabilizing | Stabilizing | Destabilizing | Destabilizing | Destabilizing | Destabilizing | Destabilizing |
| Q289L | Stabilizing | Stabilizing | Stabilizing | Destabilizing | Stabilizing | Stabilizing | Stabilizing | Stabilizing | Stabilizing |
| A261T/L285I | Destabilizing | Stabilizing | Neutral | Destabilizing | Destabilizing[a] | Destabilizing[a] | Destabilizing[a] | Destabilizing[a] | Destabilizing |
| A261S/L285I | Stabilizing | Stabilizing | Destabilizing | Destabilizing | Destabilizing[b] | Destabilizing[b] | Destabilizing[b] | Destabilizing[b] | Destabilizing |
| A261S/L285V | Destabilizing | Stabilizing | Destabilizing | Destabilizing | Destabilizing[b] | Destabilizing[b] | Destabilizing[b] | Destabilizing[b] | Destabilizing |
| A261T/L285I/Q289L | Stabilizing | Destabilizing | Neutral | Destabilizing | n/a | n/a | n/a | n/a | Destabilizing |
| A261T/L285V/Q289L | Destabilizing | Destabilizing | Neutral | Destabilizing | n/a | n/a | n/a | n/a | Destabilizing |

[a]A261T as control. [b]A261S as control. n/a, data not shown due to large uncertainty by setting double mutations as control. IMPDH, inosine monophosphate dehydrogenase.

**Supplementary Table S19: Predicted binding scores between wild-type and IMPDH mutants as well as mutation effects on binding affinity with MPA.**

| Model | CB-DOCK2 score | CSM-lig | PRODIGY-lig | PremPLI | mCSM-lig |
|---|---|---|---|---|---|
| Wild type | -7·4 | 1·9 | -5·49 | - | - |
| H108Y | -7·4 | 1·9 | -5·49 | Destabilizing | Destabilizing |
| A261T | -7·3 | 1·9 | -5·49 | Destabilizing | Destabilizing |
| A261S | -7·3 | 1·9 | -5·50 | Destabilizing | Destabilizing |
| L285I | -7·3 | 1·9 | -5·50 | Destabilizing | Destabilizing |
| L285V | -7·3 | 1·9 | -5·50 | Destabilizing | Destabilizing |
| Q289L | -7·4 | 1·9 | -5·49 | Destabilizing | Stabilizing |
| A261T/L285I | -7·3 | 1·9 | -5·50 | Destabilizing[a] | Destabilizing[a] |
| A261S/L285I | -7·3 | 1·9 | -5·50 | Destabilizing[b] | Destabilizing[b] |
| A261S/L285V | -7·3 | 1·9 | -5·50 | Destabilizing[b] | Destabilizing[b] |
| A261T/L285I/Q289L | -7·3 | 1·9 | -5·49 | n/a | n/a |
| A261T/L285V/Q289L | -7·3 | 1·9 | -5·49 | n/a | n/a |

[a]A261T as control. [b]A261S as control. n/a, data not available or not shown due to large uncertainty by setting double mutations as control. IMPDH, inosine monophosphate dehydrogenase.

**Supplementary Table S20: Reports of *Pneumocystis* pneumonia outbreaks in solid organ transplant recipients**

| First author (reference no.) | Year of report | Country | Year of first case | No. of patients | Transplanted organs | Total no. of patients from separate outbreaks starting in the same years[a] |
|---|---|---|---|---|---|---|
| Hardy[53] | 1984 | USA | 1981 | 14 | Kidney | 14 |
| Olsson[54] | 2001 | Sweden | 1987 | 5 | Kidney | 5 |
| Olsson[54] | 2001 | Sweden | 1989 | 7 | Kidney | 7 |
| Hennequin[55] | 1995 | France | 1991 | 7 | Kidney | 10 |
| Hocker[56] | 2005 | Germany | 1991 | 3 | Kidney | |
| **Neofytos[57]** | **2018** | **Switzerland** | **2005** | **5** | **Kidney** | 80 |
| **Yazaki[58]** | **2009** | **Japan** | **2005** | **8** | **Kidney** | |
| **Schmoldt[23]** | **2008** | **Germany** | **2005** | **16** | **Kidney** | |
| Pliquett[59] | 2012 | Germany | 2005 | 29 | Kidney | |
| de Boer[60] | 2007 | Netherlands | 2005 | 22 | Kidney | |
| Arichi[61] | 2009 | Japan | 2006 | 9 | Kidney | 9 |
| **Rostvet[12]** | **2013** | **Denmark** | **2007** | **29[b]** | **Kidney, liver** | 56 |
| **Gianella[62]** | **2010** | **Switzerland** | **2007** | **20** | **Kidney** | |
| Chandola[63] | 2014 | India | 2007 | 7 | Kidney | |
| Phipps[64] | 2011 | Australia | 2008 | 14 | Kidney | 84 |
| Nevez[65] | 2018 | France | 2008 | 22 | Kidney | |
| Wynckel[66] | 2011 | France | 2008 | 17 | Kidney | |
| Pliquett[59] | 2012 | Germany | 2008 | 10 | Kidney | |
| Thomas[67] | 2011 | UK | 2008 | 21 | Kidney | |
| **Neofytos[57]** | **2018** | **Switzerland** | **2009** | **31** | **Kidney** | 70 |
| Miguel Montanes[68] | 2018 | France | 2009 | 15 | Liver | |
| Thomas[67] | 2011 | UK | 2009 | 11 | Kidney | |
| Debourgogne[69] | 2014 | France | 2009 | 13 | Kidney | |
| Chapman[70] & Nankivell[71] | 2013 | Australia | 2010 | 95 | Kidney, liver, heart, lung, pancreas | 122 |
| Alanio[72] | 2017 | Belgium | 2010 | 5 | Kidney | |
| Brunot[73] | 2012 | France | 2010 | 9 | Kidney | |
| Alanio[72] | 2017 | France | 2010 | 13 | Kidney | |
| **Goto[22]** | **2017** | **Japan** | **2011** | **3** | **Kidney** | 52 |
| Mulpuru[74] | 2016 | Canada | 2011 | 10 | Kidney | |
| Gits-Muselli[75] | 2015 | France | 2011 | 14 | Kidney | |
| Urabe[76] | 2016 | Japan | 2011 | 8 | Kidney | |
| Ricci[77] | 2018 | Brazil | 2011 | 17 | Kidney | |
| Inkster[78] | 2017 | UK | 2012 | 14 | Kidney | 14 |
| **Goto[22]** | **2017** | **Japan** | **2013** | **11** | **Kidney** | 17 |
| Veronese[79] | 2018 | Italy | 2013 | 6 | Heart | |
| Desoubeaux[80] | 2016 | France | 2014 | 4 | Liver | 46 |
| Wintenberger[81] & Charpentier[82] | 2017 | France | 2014 | 10 | Lung, kidney, heart, liver | |
| Hosseini-Moghaddam[83] | 2020 | France | 2014 | 10 | Heart, kidney, liver | |
| Inkster[78] | 2017 | UK | 2014 | 11 | Kidney | |
| Alanio[72] | 2017 | UK | 2014 | 2 | Kidney | |
| McClarey[84] | 2019 | UK | 2014 | 9 | Kidney | |
| Vindrios[85] | 2017 | France | 2015 | 7 | Heart | 7 |
| **Azar[4]** | **2022** | **USA** | **2019** | **19** | **Kidney** | 19 |

Reports describing multiple separarate outbreaks are listed repeatedly according to the start year of each outbreak unless insufficient information prevents classification, e.g. in references.[70-72] Some outbreaks were included in multiple reports. Nine outbreaks in five countries investigated in this study (with a subset of samples available) are shaded according to the color scheme shown in Figure 5.
[a]Corresponding to the number of cases shown in Figure 5.
[b]First case occurred in 2007 but the first case with *impdh* mutations was detected in 2009.

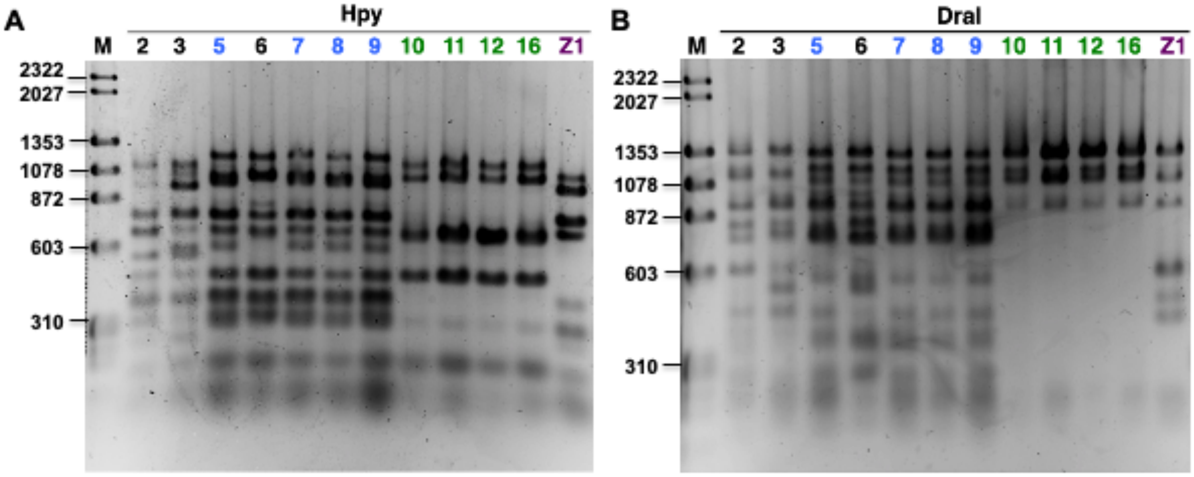

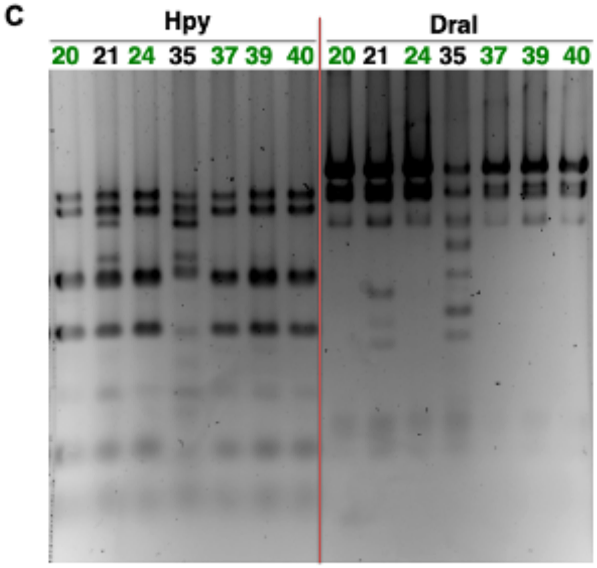

D

| Sample ID | RFLP | MLST profile | *impdh* |
|---|---|---|---|
| 5, 7, 8, 9 | Blue | 2-1-1-32-17 | H108Y |
| 10, 11, 12, 16, 20, 24, 37, 39, 40 | Green | 1-2-7-1-1 | A261T |
| 21 | Unique | 1-2-7-1-1+ | A261T |
| 2 | Unique | 1-1-1-58-1 | *impdh-3* |
| 3 | Unique | 1-1-8-37-1 | *impdh-6* |
| 6 | Unique | 1-1-8-34-6 | *impdh-7* |
| 35 | Unique | n-2-8-26-1 | *impdh-9* |
| Z1 | Unique | 1-2-1-18-53 | A261T |

**Supplementary Figure S1. Restriction fragment length polymorphism (RFLP) analysis of *P. jirovecii* samples from outbreaks in Bern and Zurich, Switzerland**. (A-C) Images of agarose gels used to separate PCR products after digestion with restriction enzymes as indicate at the top and stained with SYBR green. Numbers above each lane represent individual samples, with 2 to 40 (non-consecutive) corresponding to SB2 to SB40 from Bern and Z1 corresponding to SZ01 from Zurich in Supplementary Table S13. Samples 2 and 3 are from control patients and all others are from renal transplant patients. Blue and green colors mark two distinct groups of samples, each group sharing an identical RFLP pattern with each enzyme. Each of all other samples shows a unique pattern. DNA size markers are indicated on the left (in bp). (D) Summary of RFLP results compared with *impdh* variants and MLST profiles (Supplementary Table S13). "+' indicates coinfection with multiple strain(s) in sample 21, with the dominant strain identical to the strain for the group of 9 samples indicated in green. *impdh* variants 3, 6, 7 and 9 do not contain nonsynonymous mutations.

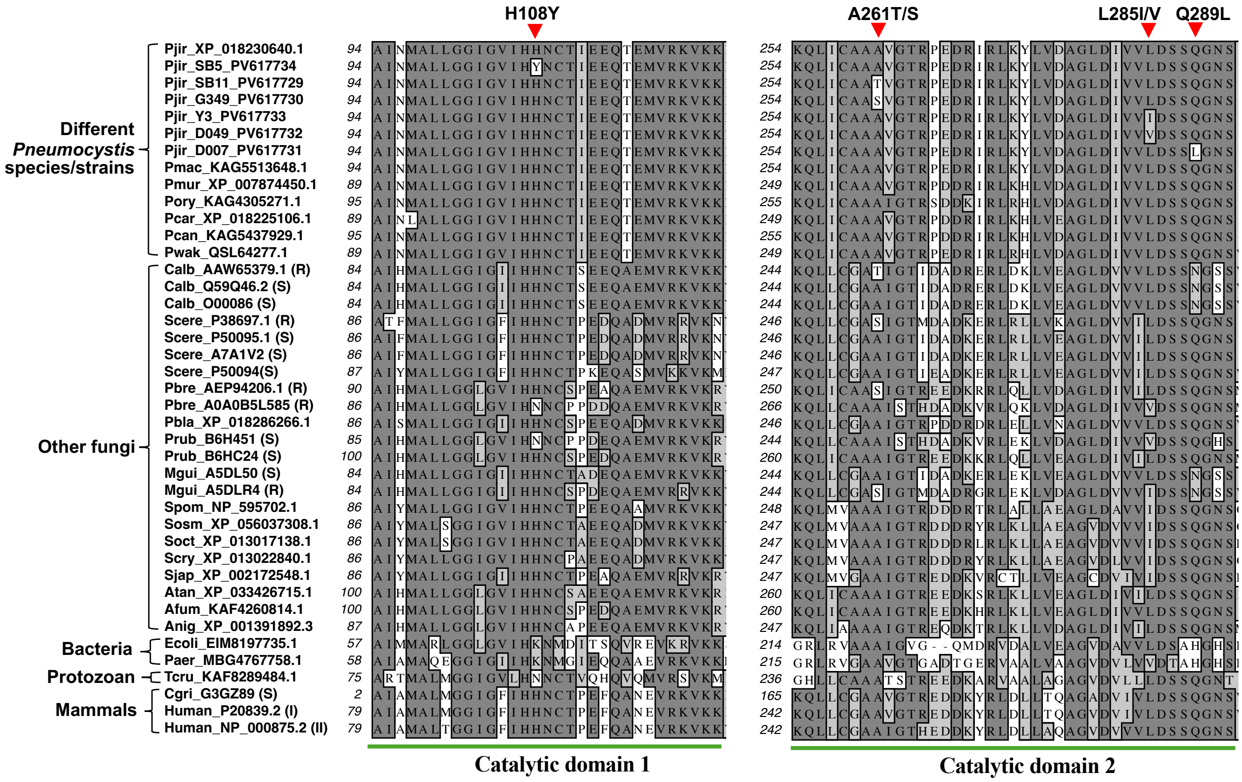

**Supplementary Figure S2: Partial alignment of inosine monophosphate dehydrogenase (IMPDH) protein sequences from *Pneumocystis* species and other organisms.** The regions shown correspond to the amino acid residues 94-130 (left panel) and 254-292 (right panel) of the IMPDH protein (Pjir_XP_018230640.1 on the top) in the *P. jirovecii* RU7 reference genome (NCBI RefSeq assembly no. GCF_001477535.1), which is designated as the wild type (IMPDH variant 1). The sequences from the second to seventh lines are from six strains of *P. jirovecii* (denoted as Pjir) containing six different *impdh* mutations at four amino acid positions as indicated by red triangles (including two distinct mutations each at 261 and 285). Both regions are located within the catalytic domains of the IMPDH enzyme based on reference.[86] NCBI Protein database accession numbers are indicated on the left after the abbreviated species or strain names, with S and R in parentheses representing fungal species known to be susceptible and resistant to mycophenolic acid, respectively. Pjir, *P. jirovecii*; Pmac, *P. macacae*, Pmur, *P. murina*; Pory, *P. oryctolagi*, Pcar, *P. carinii*, Pcan, *P. canis*, Pwak, *P. wakefieldiae*; Calb, *Candida albicans*; Scere, *Saccharomyces cerevisiae*; Pbre, *Penicillium brevicompactum*; Pbla, *Phycomyces blakesleeanus*; Mgui, *Meyerozyma guilliermondii*; Spom, *Schizosaccharomyces pombe*; Sosm, *S. osmophilus*; Soct, *S. octosporus*; Scry, *S. cryophilus*; Sjap, *S. japonicus* : Atan, *Aspergillus tanneri*; Afum, *A. fumigatus*; Anig, *A. niger*; Ecoli, *Escherichia coli*; Paer, *Pseudomonas aeruginosa*; Tcru, *Trypanosoma cruzi*; Cgri, *Cricetulus griseus* (Chinese hamster).

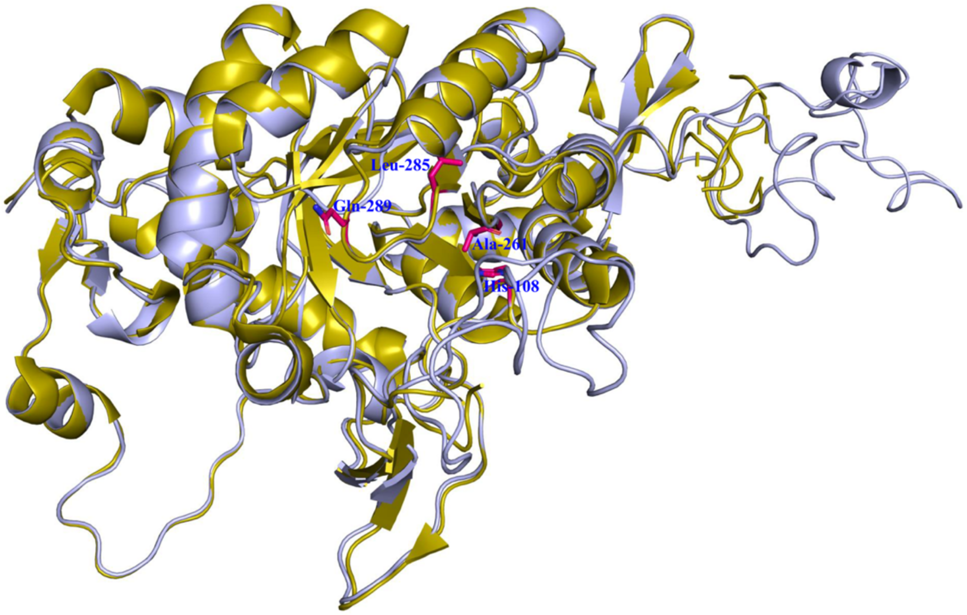

**Supplementary  Figure S3: Comparison of the modeled wild-type *P. jirovecii* IMPDH structure with chain A of the reference IMPDH structure 1JR1**.
Inosine monophosphate dehydrogenase (IMPDH) structure is colored in light blue, and 1JR1 in olive. Highlighted in pink are the four residues that are prone to mutation in transplant patients receiving mycophenolic acid.

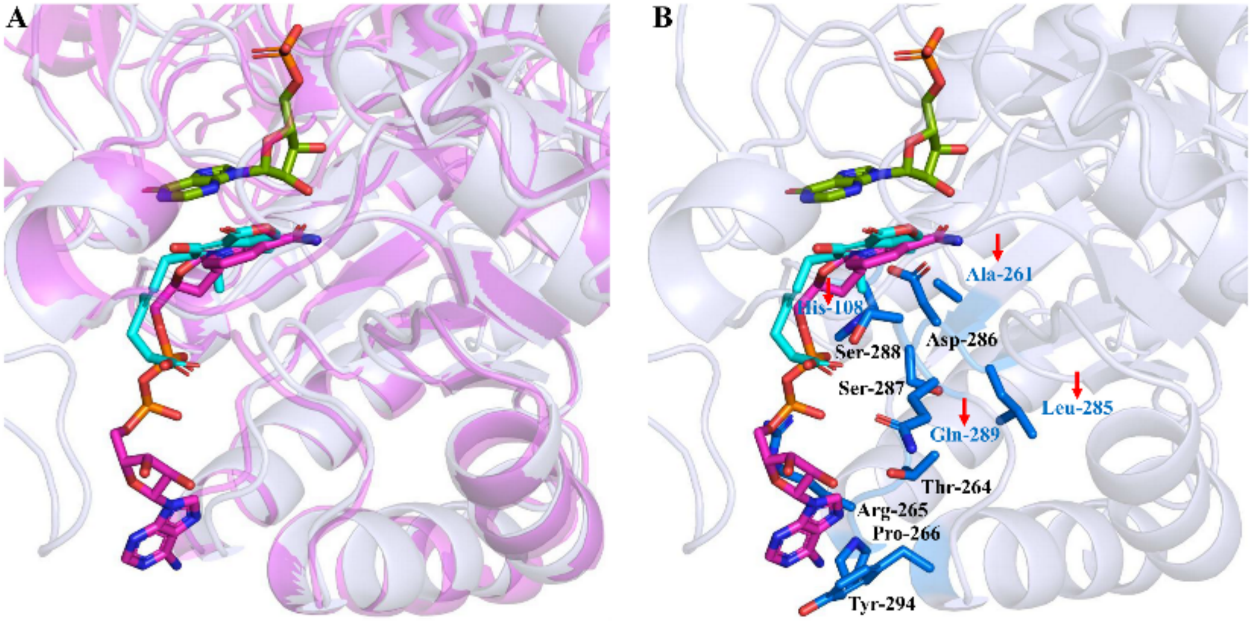

**Supplementary  Figure S4: Alignment of the wild-type docking model to human IMPDH structure with the PDB ID of 6U8N**. (**A**) Alignment between wild-type docking model and 6U8N, where the docking model is colored in light blue (MPA: cyan by element), and 6U8N is colored in magenta ($NAD^+$: magenta by element; IMP: split pea by element); (**B**) Representative key residues around $NAD^+$ within a 5Å radius around MPA are highlighted and four high-frequency mutation residues are pointed by red arrows.
